# Machine Learning Prediction of Food Processing

**DOI:** 10.1101/2021.05.22.21257615

**Authors:** Giulia Menichetti, Babak Ravandi, Dariush Mozaffarian, Albert-László Barabási

## Abstract

Despite the accumulating evidence that increased consumption of ultra-processed food has adverse health implications, it remains difficult to decide what constitutes processed food. Indeed, the current processing-based classification of food has limited coverage and does not differentiate between degrees of processing, hindering consumer choices and slowing research on the health implications of processed food. Here we introduce a machine learning algorithm that accurately predicts the degree of processing for any food, indicating that over 73% of the U.S. food supply is ultra-processed. We show that the increased reliance of an individual’s diet on ultra-processed food correlates with higher risk of metabolic syndrome, diabetes, angina, elevated blood pressure and biological age, and reduces the bio-availability of vitamins. Finally, we find that replacing foods with less processed alternatives can significantly reduce the health implications of ultra-processed food, suggesting that access to information on the degree of processing, currently unavailable to consumers, could improve population health.

## Introduction

Unhealthy diet is a major risk factor for multiple non-communicable diseases, from obesity and type 2 diabetes to coronary heart disease (CHD) and cancer, together accounting for 70% of mortality and 58% of morbidity worldwide [1,2]. Traditionally, consumers rely on food category-based dietary recommendations like the Food Pyramid (1992 [3]) or MyPlate (2011 [4]), which define the mix of fruits, vegetables, grains, dairy and protein-based foods that constitute a healthy diet. In recent years, however, an increasing number of research studies and dietary guidelines have identified the important role and separate health effects of food processing [5–10]. Observational studies, meta-analysis, and controlled metabolic studies suggest that dietary patterns relying on unprocessed foods are more protective than the processing-heavy Western diet against disease risk [11, 12]. The role of processed food has reached food policy and is now embodied in several expertise-based food classification systems used in cohort studies such as EPIC [13, 14], and led to the expansion of food description and ontology systems such as LanguaL [15], FoodEx2 [16–19], and FoodOn [20, 21].

NOVA [22–24] is a classification system for food processing, widely used in epidemiological studies. It categorizes individual foods into four broad categories: *unprocessed or minimally processed* (NOVA 1), like fresh, dry or frozen fruits or vegetables, grains, legumes, meat, fish and milk; *processed culinary ingredients* (NOVA 2), like table sugars, oils, fats and salt; *processed foods* (NOVA 3), like canned food, simple bread and cheese; and *ultra-processed products* (NOVA 4), industrial formulations typically of five or more ingredients including substances not commonly used in culinary preparations, such as additives whose purpose is to imitate sensory qualities of fresh food. Examples of ultra-processed foods include packaged breads, cookies, sweetened breakfast cereals, margarines, sauces and spreads, carbonated drinks, hot dogs, hamburgers and pizzas.

Epidemiological studies have documented significant associations between greater consumption of the highest NOVA category of processing (NOVA 4, ultra-processed food) and diseases onset [25–31], including links to obesity [32], CHD [30, 33], diabetes mellitus [34, 35], cancer [29, 36], and depression [28]. Finally, a randomized controlled metabolic trial in 20 adults has confirmed the short-term adverse effects on caloric intake and weight gain of ultra-processed foods [31].

Despite its success, NOVA is purely descriptive in nature, leading to a laborious and incomplete classification task, limiting research into the impact of processed food [37–39]. First, NOVA relies on expertise-based manual evaluation of each food, and assigns a unique class to only 35% of the foods catalogued by the U.S. Department of Agriculture (USDA), decomposing the remaining part of the database in ingredients to be further analyzed (SI Section 1). Second, the classification of composite recipes, products, and mixed meals including different types of processed items – a large proportion of the food supply – is not straightforward. Whilst, an accurate ingredient list of all complex foods would allow for an ingredient-based classification, a full and quantified ingredient list is seldom, if ever, available to the consumer, and when available, it shows considerable variability from database to database. Interestingly, even in presence of detailed ingredient information, the consistency across nutrition specialists in assigning NOVA classes was found to be low [39]. Given these data limitations, current approaches have classified all foods with at least one ultra-processed ingredient as ultra-processed [29, 40], reducing the discrimination of such foods in relation to health outcomes [41, 42]. Third, all the observed risk for the NOVA classification is in the NOVA 4 class, representing a large and heterogeneous category of ultra-processed food that limits our ability to investigate the health implications of different gradations of processing. Indeed, according to NOVA, many nations derive up to 60% or more of their average caloric intake from ultra-processed foods [42–44]. While NOVA allows for a more refined analysis, all foods within this class are considered to have identical health consequences. These perceived homogeneity of NOVA 4 foods limits both scientific research and practical consumer guidance on the health effects of differing degrees of processing. It also reduces the industry’s incentives to reformulate foods towards less processed offerings, shifting investments from the ultra-processed NOVA 4 foods to the less processed NOVA 1 and NOVA 3 categories. To overcome these limitations, here we introduce FoodProX, a machine learning classifier that takes as input nutritional measures, and is trained to predict the degree of processing of any food in a reproducible, portable, and scalable fashion. We rely on nutrients as input because: 1) The list of nutrients in a food is consistently regulated and reported worldwide; 2) Their quantities in unprocessed food are constrained by physiological ranges determined by biochemistry [45]; 3) Food processing systematically and reproducibly alters nutrient concentrations through combinatorial changes detectable by machine learning (Figures 1A-B). FoodProX allows us to define a continuous index (FPro) that captures the degree of processing of any food, and helps us quantify the overall diet quality of individuals, ultimately unveiling the statistical correlations between the degree of processing characterizing individual diets and multiple disease phenotypes.

**Figure 1:**
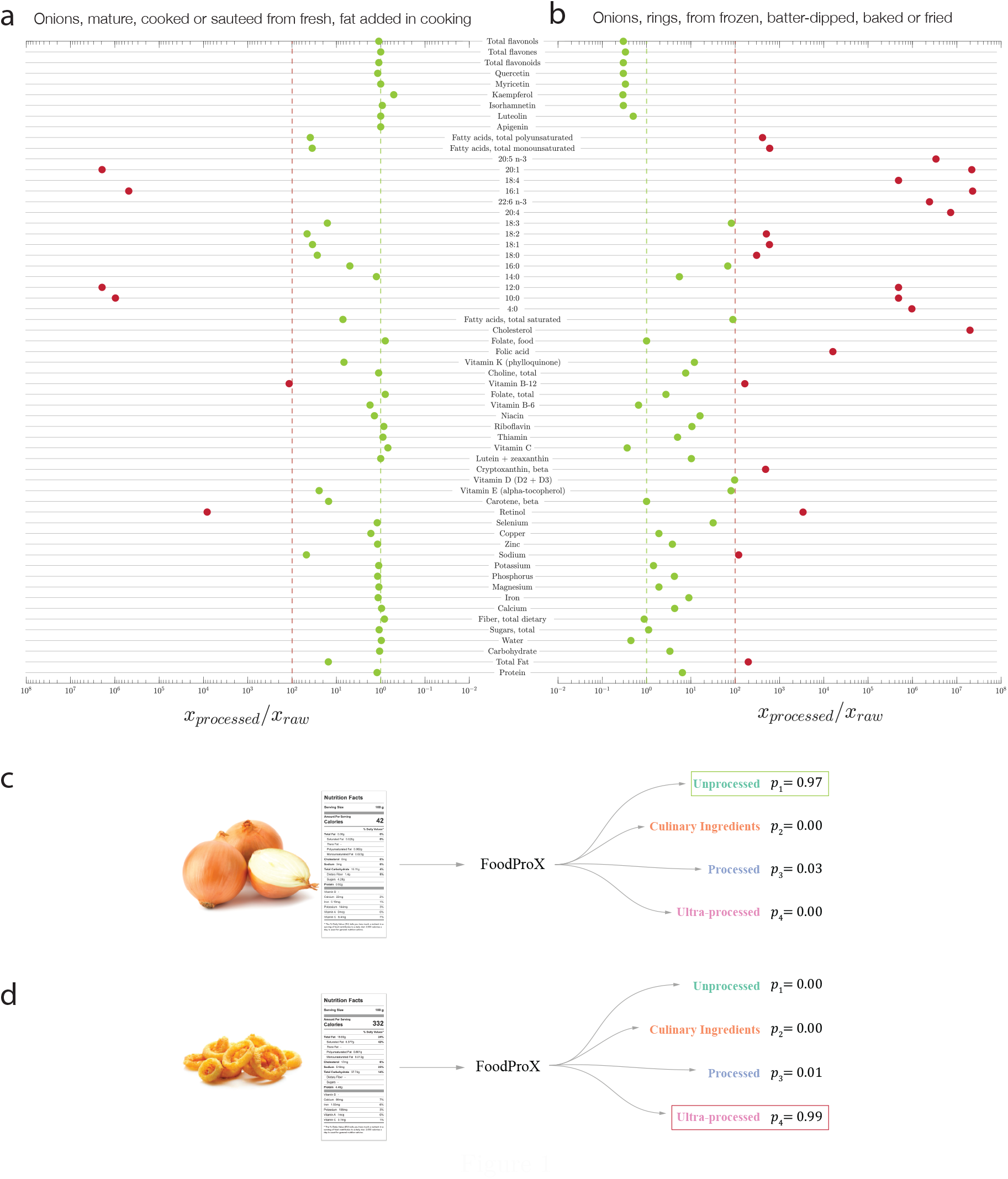
Food Processing and Nutrient Changes (FoodProX). **(a)-(b)** Ratio of nutrient concentrations for 100 g of Sauteed Onion and Onion Rings compared to Raw Onion, indicating how processing alters the concentration of multiple nutrients. All nutrients in excess compared to the concentrations found in the raw ingredient are shown in red. **(c)-(d)** We trained FoodProX, a random forest classifier over the nutrient concentrations within 100 g of each food, tasking it to predict its processing level according to NOVA. FoodProX represents each food by a vector of probabilities *{p*_*i*_*}*, capturing the likelihood of being classified as unprocessed (NOVA 1), processed culinary ingredients (NOVA 2), processed (NOVA 3), and ultra-processed (NOVA 4). The highest probability determines the final classification label, highlighted in a box on the right.

## Results

The manual procedure behind NOVA, relying on the ingredients of food, has allowed a straightforward labeling to 2,484 foods reported in the National Health and Nutrition Examination Survey (NHANES) 2009-2010, representing 34.25% of items consumed by a representative sample of the U.S. population [26] (SI Section 1). The remaining foods are either not classified, or require further decomposition into their differing food ingredients, often not reported by the manufacturers. In contrast, the basic nutrient profile of foods and beverages is always disclosed, mandated by law in most countries. For example, the USDA Standard Reference database (USDA SR Legacy), catalogues the nutrient profile of 7,793 foods with resolutions ranging from 8 to 138 nutrients (Figure S1) [46], and USDA FNDDS reports between 65 to 102 nutrients for all foods consumed by NHANES participants [47, 48].

Our work relies on the hypothesis that the nutrient profiles of unprocessed or minimally processed foods are generally constrained within common physiologic ranges [45]. The nutrient profile can be widely altered by the physical, biological and chemical processes involved in food preparation and conservation, thus correlating with the degree of processing undertaken. Indeed, among the nutrients reported in raw onion, 3/4 change their concentration in excess of 10% in the fried and battered version of the product, and more than half by 10-fold (Figure 1B). We lack however, a single nutrient “biomarker” that accurately tracks the degree of processing; instead we observe changes in the concentration of multiple nutrients, whose combinations jointly correlate with processing. This complexity advocates for the use of machine learning, designed to capture the combinatorial explosion of nutrient alterations.

### FoodProX Algorithm

To train FoodProX, a multi-class random forest classifier, we leveraged the nutrient concentrations provided in FNDDS 2009-2010 for the foods classified in Ref. [26] (Figure 1D). FoodProX takes as input the list of nutrients in any food and offers as output a vector of four probabilities {*p*_*i*_}, representing the likelihood that the respective food is classified as unprocessed (*p*_1_, NOVA 1), processed culinary ingredients (*p*_2_, NOVA 2), processed (*p*_3_, NOVA 3), and ultra-processed (*p*_4_, NOVA 4). The highest of the four probabilities determines the final classification label for each food (Figure 1C-D).

As 90% of foods in USDA Branded Food Products Database report less than 17 nutrients, we also tested the algorithm’s predictive power with the 12 gram-based nutrients mandated by FDA [49] (SI Section 1). To evaluate the performance of FoodProX we measure the Area Under the Receiver Operating Characteristic (AUC), defined as the probability that a random sample from the class of interest will have a higher score than a random sample from any other class. We identified consistently high AUC values for all the considered nutrient subsets: 0.9804±0.0012 for NOVA 1, 0.9632±0.0024 for NOVA 2, 0.9696±0.0018 for NOVA 3, and 0.9789±0.0015 for NOVA 4, significantly far from a random performance with AUC=0.5, describing a model with no discriminating power (see SI Section 2.1, Figure S5, and Table S6 for a detailed analysis of the cross-validated performances, including precision and recall).These results confirm that changes in nutrient content have remarkable predictive power in capturing the extent of food processing. Furthermore, we find that no single nutrient drives the predictions, but the predictive signal is rooted in combinations of changes spanning multiple nutrients (see SI Section 2.2 for a detailed permutation feature importance and Shapley value analysis [50]).

We visualize the decision space of the classifier by performing a principal component analysis over the probabilities {*p*_*i*_}, observing that the list of foods manually classified by NOVA is limited to the three corners of the phase space, to which the classifier assigns dominating probabilities (Figure 2A). We used FoodProX to classify all foods and beverages that lacked prior manual NOVA classification in FNDDS (65.75% of total). We found that 7.39% of FNDDS consists of NOVA 1; 0.90%, NOVA 2; 18.36%, NOVA 3; and 73.35%, NOVA 4 foods (Figure 2B). Yet, many previously unclassified foods are often inside the phase space, indicating that they lack a dominating probability, hence the assignment of a single NOVA class is somewhat arbitrary (Figure 2B). The detection of this ambiguity is a strength of FoodProX, reflecting the observation that a four-class classification encodes a great extent of nutrient variability associated with different food processing methods and intensities [51]. For example, while the classifier confidently assigns “Raw Onion” to NOVA 1 (*p*_1_ = 0.9651), and “Onion rings prepared from frozen” (*p*_4_ = 0.9921) to NOVA 4, it accurately offers an intermediate confidence for “Onion, Sauteed,” placing it with probability *p*_4_ = 0.6521 in NOVA 4, and with probability *p*_3_ = 0.2488 in NOVA 3.

**Figure 2:**
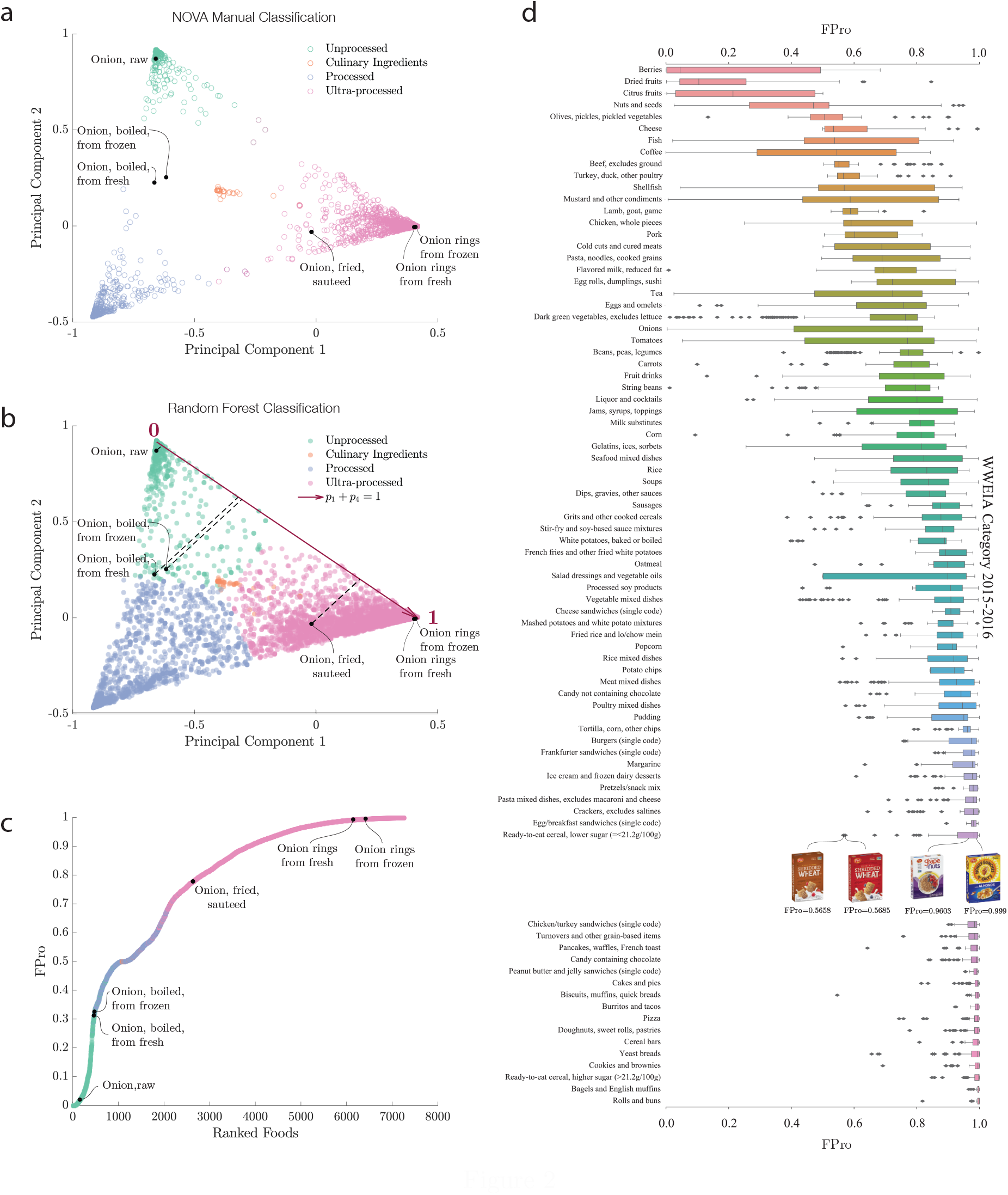
NOVA Classification and Processing Score. **(a)** Visualization of the decision space of FoodProX via principal component analysis of the probabilities *{p*_*i*_*}*. The manual 4-level NOVA classification assigns unique labels to only 34.25% of the foods listed in FNDDS 2009-2010 (empty circles). The classification of the remaining foods remains unknown (“Not Classified”), or must be further decomposed into ingredients (“Composite Recipe”). The list of foods manually classified by NOVA is largely limited to the three corners of the phase space, foods to which the classifier assigns dominating probabilities. **(b)** FoodProX assigned NOVA labels to all foods in FNDDS 2009-2010. The symbols at the boundary regions indicates that for these foods the algorithm’s confidence in the classification is not high, hence a 4-class classification does not capture the degree of processing characterizing that food. For each food *k*, the processing score *FPro*_*k*_ represents the orthogonal projection (black dashed lines) of 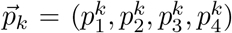 onto the line *p*_1_ + *p*_4_ = 1 (highlighted in dark red). **(c)** We ranked all foods in FNDDS 2009/2010 according to *FPro*. The measure sorts onion products in increasing order of processing, from “Onion, Raw”, to “Onion rings, from frozen”. **(d)** Distribution of *FPro* for a selection of the 155 Food Categories in What We Eat in America (WWEIA) 2015-2016 with at least 20 items (SI Section 2). WWEIA categories group together foods and beverages with similar usage and nutrient content in the U.S. food supply [52]. All categories are ranked in increasing order of median *FPro*, indicating that within each food group, we have remarkable variability in *FPro*, confirming the presence of different degrees of processing. We illustrate this through four ready-to-eat cereals, all manually classified as NOVA 4, yet with rather different *FPro*. While the differences in the nutrient content of Post Shredded Wheat ‘n Bran (*FPro* = 0.5658) and Post Shredded Wheat (*FPro* = 0.5685) are minimal, with lower fiber content for the latter, the fortification with vitamins, minerals, and the addition of sugar, significantly increases the processing of Post Grape-Nuts (*FPro* = 0.9603), and the further addition of fats results in an even higher processing score for Post Honey Bunches of Oats with Almonds (*FPro* = 0.9999), showing how *FPro* ranks the progressive changes in nutrient content.

### Food Processing Score (FPro)

The observation that enforcing discrete classes causes inherent challenges in food classification prompted us to introduce the Food Processing Score (*FPro*), a continuous variable with *FPro* = 0 for raw ingredients, and *FPro* → 1 for ultra-processed foods. We define the *FPro* of a food *k* as

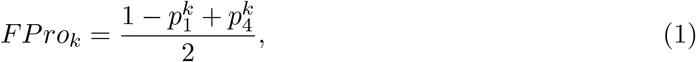

capturing the progressive changes in nutrient content induced by processing. In other words, the FPro score measures the trade-off between the confidence that the FProX algorithm has in classifying the food item *k* as NOVA 1 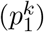 or NOVA 4 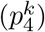. For example, *FPro* incrementally increases from raw onion (*FPro* = 0.0203) to boiled onion (*FPro* = 0.3126), fried onion (*FPro* = 0.7779), and onion rings from frozen ingredients (*FPro* = 0.9955, Figure 2C). *FPro* allows us to unveil the degree of processing characterizing different food preparation techniques, assigning lower values to foods made from fresh ingredients than those made from frozen ingredients (Figure 2C). *FPro* also permits classification of complex recipes and mixed dishes, identifying the overall average processing of the item or meal.

As noted above, nearly 20% of the U.S. diet comprises foods in NOVA 3, and over 70%, in NOVA 4. The observed variability of *FPro* for foods that belong in the same NOVA class prompted us to analyze the variations in degree of processing within subgroups of foods and beverages, as captured by What We Eat in America (WWEIA) categorization [52]. We find that different food products within the same NOVA classification and WWEIA categories show remarkable variability in *FPro*, confirming the presence of different processing fingerprints within each category (Figure 2D). For example, if we select four products from the same brand (Post) of breakfast cereals that are all manually classified as NOVA 4 (i.e., considered identically processed), we observe that *FPro* differentiate them and captures progressive alterations in fiber content, fortification with vitamins and minerals, and addition of sugar and fats (Figure 2D, insert, and Section S2.5).

### Stability and Robustness of FPro

Real world foods vary in nutritional content, affected by recipe variations, production methods, soil quality, storage time, and changing government regulations [53]. This degree of variability, coupled by measurement and reporting uncertainties, raises a fundamental question: is FPro robust against the expected variability and uncertainty in nutrient content? To address this, we first explored how nutritional values for the same food change through different FNDDS cycles, focusing on the 5,632 foods whose nutrient profile is reported both in FNDDS 2009-2010 and 2015-2016 (SI Section 7). For example, in the less processed category, for “Milk, calcium fortified, cow’s, fluid, whole” we find that 14 nutrients have changed between 2009 and 2015, including a 3.65 fold decrease in Calcium content. In a similar fashion, the highly ultra-processed “Cookie, vanilla with caramel, coconut, and chocolate coating” shows variation in 46 nutrients, with 6 fold decrease in monounsaturated fatty acid 20:1. Despite such significant changes in the content of some nutrients, FPro shows remarkable stability: milk’s FPro goes from 0.0010 to 0.0011, consistently classifying it as unprocessed, and for the cookie FPro goes from 0.9943 to 0.9965, staying firmly in the ultra-processed category. Overall, for foods whose nutrients maximally change between 10% and 50% of their original value, we observe an absolute shift in FPro of 0.001556 (quartiles *Q*_1_(25%)=0.000222, *Q*_3_(75%)= 0.004764). Allowing up to 1000% of nutrient variability does not significantly alter our findings, since the expected change of FPro per food reaches 0.003312 (quartiles *Q*_1_(25%)= 0.000722, *Q*_3_(75%)= 0.011310). The observed FPro stability is rooted in the fact that FPro’s value is driven by the nutrient panel as a whole, and not by the concentration of any single nutrient (see SI Sections S7, for further details on data sampling and variability).

Currently, the chemical information available to train FPro does not track the concentration of additives as they are rarely available for most foods. Yet, additives like tertiary butylhydroquinone, acetylated monoglycerides, polysorbates, sodium stearoyl lactylate and sodium aluminum phosphate, represent obvious signatures of food processing [54], raising the question on how much improvement in predictive power we could obtain if information on additives would be available. We therefore relied on the Open Food Facts, that compiles an extensive list of food additives, including artificial colors, artificial flavors, and emulsifiers [55], to test the FPro’s ability to absorb information on additives. From the Open Food Facts website we collected 233,831 nutritional records, annotated with NOVA labels according to a heuristic described in [56]. We then trained and validated two models: (1) FProX leveraging only nutrition facts as introduced before, (2) a modified FProX, using nutrition facts and the available information on the number of additives (SI Section 6). While model (2) displays slightly better performance, with AUC 0.9926±0.0003 for NOVA 1, 0.9878±0.0047 for NOVA 2, 0.9653±0.0010 for NOVA 3, 0.9782±0.0007 for NOVA 4, we find that the performance of model (1) is largely indistinguishable, reaching AUC 0.9880±0.0006 for NOVA 1, 0.9860±0.0045 for NOVA 2, 0.9320±0.0015 for NOVA 3, 0.9508±0.0009 for NOVA 4. In other words, while information on additives can improve FProX’s performance, changes in the nutrient profile already carry the bulk of the predictive power. This aspect is further confirmed by the performance of a classifier purely based on the number of additives: in this scenario the number of false positives for NOVA 1, 2, and 3 remarkably increases, affecting the precision of the model, and showing predictive power only for NOVA 4 (see SI Section 6 for a detailed comparison of the models).

Finally, we investigate how FPro, a measure of the nutritional quality of food, changes depending on where the food was prepared, exploring if it can distinguish between home-cooked food, food prepared in stores, canteens, restaurants, fast foods, and products available in vending machines. Leveraging data from NHANES, we ranked the 10 most popular food sources in increasing value of FPro. The lowest FPro is observed for “Grown or caught by you and someone you know” (median FPro=0.4423) and “Residential dining facility” (median FPro=0.6238), confirming the less processed nature of home prepared foods. We obtain a much higher FPro for “Restaurant fast food/pizza” (median FPro=0.9060) and “Vending machine” (median FPro=0.9800), confirming its reliance on ultra-processed ingredients. This result indicates that FPro can differentiate food sources, styles and quality of food preparation (see Section S2.6 for the statistical analysis).

### Individual Processing Score

The significant contribution of ultra-processed food to American dietary intake, and *FPro*’s ability to demonstrate heterogeneity in the extent of food processing within the broad category of ultra-processed food, prompts us to assess the contribution of processed food to the diet of each individual, jointly weighted by both the extent of processing and the contribution to caloric intake. This is provided by the individual Food Processing Score (*iFPro*),

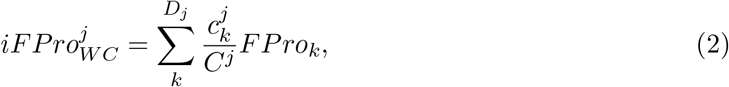

which varies between 0 and 1, where *D*_*j*_ is the number of dishes consumed by individual *j, C*^*j*^ is the daily total amount of consumed calories, and 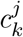 is the amount of calories contributed by each food item. A gram-based *iFPro*_*W G*_, captures the fraction of grams in a diet supplied by processed food (Eq. S6).

We calculated *iFPro* for 20,047 individuals with dietary records in a representative U.S. national sample from NHANES 1999-2006. NHANES relies on 24-hour recalls to capture dietary intake, a widely used methodology in epidemiology, with well-explored statistical characteristics [57]. Indeed, studies collecting multiple days of intake data per individual indicate that it is statistically more efficient to increase the number of individuals in the sample than increase the number of days beyond two per individual [58]. As Figure 3C shows, the median *iFPro*_*W C*_ for the American population is 0.7872, confirming a high reliance on intake on ultra-processed food [42–44]. More importantly, *iFPro* allows us to identify differential reliance on processed food. Consider individuals 𝒜 and ℬ, men of similar age (47 vs. 48 years old, Figure 3A-C), with similar number of reported daily unique dishes (12.5 vs. 13 dishes) and comparable caloric intake (2,016 vs. 1,894 kcal). Yet, the diet of individual 𝒜 has *iFPro*_*W C*_ = 0.3981, as it relies on unprocessed ingredients and home cooking. In contrast, ℬ has *iFPro*_*W C*_ = 0.9572, given his consumption of hot dogs, hamburgers, French fries, pizza, and Kit Kat (see SI Section 3.5 for a full analysis of the correlation between *iFPro*_*W C*_ and food group consumption in the US population). As we show next, these differences correlate with different health outcomes.

**Figure 3:**
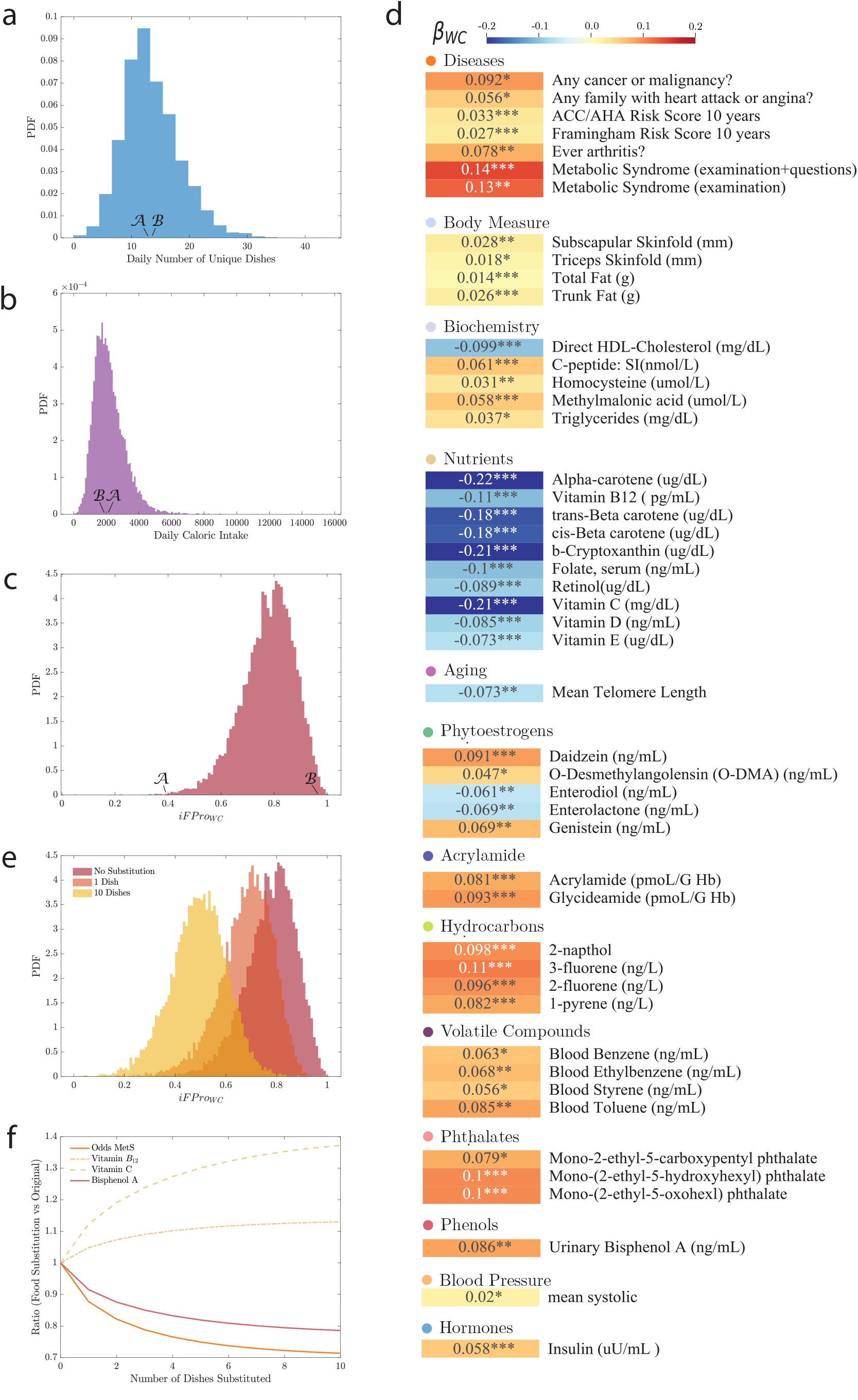
Health implications and Food Substitution. For each of the 20,047 individuals in NHANES (1999-2006), 18+ years old with dietary records [59], we calculated the individual diet processing scores *iFPro*_*W C*_. **(a)** The average number of unique dishes reported in the dietary interviews, highlighting two individuals 𝒜 and ℬ, with comparable number of dishes, 12.5 and 13 reported, respectively. **(b)** The distribution of average daily caloric intake, showing that individuals ℬ and 𝒜 have similar caloric intake of 1,894 and 2,016 kcal, respectively. **(c)** The distribution of *iFPro*_*W C*_ for NHANES, indicating that individuals 𝒜 and ℬ display significant differences in *iFPro*_*W C*_, with ℬ’s diet relying on ultra-processed food (*iFPro*_*W C*_ = 0.9572), and 𝒜 reporting simple recipes (*iFPro*_*W C*_ = 0.3981) (Figure S13). **(d)** We measured the association of various phenotypes with *iFPro*_*W C*_, correcting for age, gender, ethnicity, socio-economic status, BMI, and caloric intake (SI Section 4). We report the standardized *β* coefficient, quantifying the effect on each exposure when the Box-Cox transformed dietary scores increase of one standard deviation over the population. For continuous exposures the coefficients are fully standardized, while for logistic regression (disease phenotypes) we opted for partially standardized coefficients to help interpretability (SI Section 4). Each variable is color-coded according to *β*, positive associations shown in red, and negative associations in blue. **(e)** Changes in *iFPro*_*W C*_ when one (orange) or up to ten (yellow) dishes are substituted with their less processed versions, following the prioritization rule defined in Eq. S8. **(f)** The impact of substituting different number of dishes on the odds of metabolic syndrome, concentrations of vitamin *B*_12_, vitamin C, and bisphenol A, showing that a minimal substitution strategy can significantly alter the health implications of ultra-processed food.

### Health Implications

To quantify the degree to which the consumption of ultra-processed foods, graded across their entire range, correlate with health outcomes, we investigated exposures and phenotypes provided in NHANES 1999-2006 [59]. Inspired by [60], we performed an Environment-Wide Association Study (EWAS), an alternative to single-exposure epidemiological studies, to identify the most significant environmental factors associated with different health phenotypes in a non hypothesis-driven fashion. We measured for each variable the association with the diet processing score *iFPro*, adjusting for age, sex, ethnicity, socio-economic status, BMI and caloric intake (Figure S17). After False Discovery Rate (FDR) correction for multiple testing, 209 variables survive (SI Section 4), documenting how the consumption of ultra-processed foods relates to health.

#### Metabolic and Cardiovascular Risk

We find that individuals with a high food processing score show positive associations with risk of metabolic syndrome (MetS), diabetes (fasting glucose), and also (consistent with poor diets clustering in families) a family history of heart attack or angina, in line with earlier findings (SI Section 4) [60–62]. The increased risk of cardiovascular disease is further confirmed by the significant positive association of *iFPro* with both Framingham and ACC/AHA Risk Scores (Figure 3D) [63, 64].

Overall, individuals with a higher food processing score exhibit higher blood pressure, trunk fat and subscapular skinfold, measures of obesity, and blood insulin and triglyceride levels; and lower “good” HDL cholesterol. Further novel findings indicate a higher prevalence of type 2 diabetes (C-peptide), inflammation (C-Reactive Protein), vitamin deficiency (homocysteine, methylmalonic acid), and inflammatory arthritis [65–70]. We also find an inverse association between *iFPro* and telomere length, which can be affected by diet through inflammation and oxidation [71], suggesting a higher biological age for individuals with higher reliance on more ultra-processed foods (SI Section 4, Figure S21).

#### Vitamins and Phytoestrogens

A greater consumption of more extensively processed foods correlates with lower levels of vitamins in our blood stream, like Vitamin *B*_12_ and Vitamin C, despite the fact that ultra-processed breakfast cereals and refined flours are frequently fortified with these vitamins and minerals.

In addition, *iFPro* allows us to discriminate plant-based diets relying on legumes, whole grains, fruits and vegetables, from diets that include ultra-processed plant-based meat, dairy substitutes, and plant-based drinks [72]. For example, we observe a positive correlation between *iFPro*_*W C*_ and the urine levels of daidzein, genistein, and their bacterial metabolite o-desmethylangolensin – all bio-markers of soy, abundant in diets that rely on highly processed soy protein foods. On the other hand, enterolactone and enterodiol, gut metabolites of plant lignans (fiber-associated compounds found in many plant families and common foods, including grains, nuts, seeds, vegetables, and drinks such as tea, coffee or wine) are inversely associated with *iFPro*_*W C*_ [73].

#### Chemical Exposures

Diet rich in highly processed food shows association with increased carcinogenic, diabetic, and obesity-inducing food additives and neoformed contaminants, several of which represent previously unknown relationships. For instance, we find positive associations with acrylamide and polycyclic aromatic hydrocarbons, present in heat treated processed food products as a result of Maillard reaction, benzenes (abundant in soft drinks), furans (common in canned and jarred foods), PCBs (processed meat products), perfluorooctanoic acids and phthalates (found in the wrappers of some fast foods, microwavable popcorn, and candy), and environmental phenols like the endocrine disruptor bisphenol A (linked to plastics and resins for food packaging) [74–79]. Importantly, all these represent compounds not reported in food composition databases, but recovered in blood and urine (Figure S21).

Taken together, our ability to distinguish the degree of processing of foods in individual diets allowed us to unveil multiple correlations between diets with greater reliance on ultra-processing and health outcomes. This approach leads to results that cannot be captured with the existing NOVA classification (SI Section 4, Figures S18 and S19). Specifically, if we rely on the manual NOVA 4 and the panel of exposures in NHANES, we recover only 92 significant associations. Among the missing associations are the inverse correlation of a more ultra-processed diet with vitamin D and folate blood levels, and the positive correlation with c-peptide, homocysteine, and blood pressure (SI Section 4).

### Food Substitution

The high reliance of ultra-processed food among the U.S. population (Figure 3C) prompts us to ask: what kind of interventions could help us reduce the observed health implications? Given the challenges of large behavioral shifts [80, 81], here we assume that an individual does not need to overhaul her entire diet, but replace the most processed items she consumes with less processed versions of the same item. To minimize the dietary shifts required, we identify within each individual’s diet the item with high caloric contribution and for which there are significantly less processed alternatives (Eq. S8), preserving the broad food class (WWEIA) of the initial choice (SI Section 5). For example, we replace Kix cereals (*FPro* = 0.9998) with shredded wheat and bran cereals (*FPro* = 0.5091), and spread cheese from a pressurized can (*FPro* = 0.9648) with provolone cheese (*FPro* = 0.5001).

We find that the isocaloric replacement of a single item from the average of 19.28 food items daily consumed by U.S. adults reduces the median *iFPro*_*W C*_ by 12.15%, from an overall score of 0.7872 to 0.6915 (Figure 3E). Based on observed associations with phenotypes, this translate to a decrease in the odds of metabolic syndrome by 12.25%, a lower concentration of urinary bisphenol A by 8.47%; and an increased blood concentration of vitamin *B*_12_ and vitamin C by 4.83% and 12.31%, respectively (Figure 3F and Eqs. S9 and S10). Furthermore, the substitution of 10 food items, about half the daily reported items, leads to 37.03% decrease in *iFPro*_*W C*_, with associated changes of −21.43% for bisphenol A, +13.02% for blood vitamin *B*_12_, and +37.26% for blood vitamin C.

Overall, we find that modest substitution strategies made possible by *FPro* can preserve the general nature of an individual’s diet but reduce reliance on more highly processed food and successfully moderate the observed health implications. As a first step to implement such strategies in the real world, consumers and policymakers must be empowered with information on the degree of processing characterizing the foods in their food environment available to consume, and their potential alternatives. To successfully and sustainably improve population diets, interventions will need to combine individual nudges and behavior change strategies with interventions that more precisely address the ecological, system-level phenomena that people are exposed to, including addressing major gaps and disparities in life-style and healthy food access (i.e., nutrition security).

## Discussion

In this paper, we introduced FPro, a continuous processing score combining in a non-linear fashion features of processing techniques learned from the NOVA manual labels, with nutrient concentrations from food composition data. FPro is derived from FoodProX, a classifier that shows a remarkable ability to replicate the manual NOVA classification from nutritional information, confirming that NOVA classes result in distinct patterns of nutrient alterations, accurately detected by machine learning. Importantly, FoodProX allows us to build upon and extend the current NOVA classification in several crucial respects, offering automated and reproducible classification of foods across multiple national and commercial databases, ability to classify complex recipes and mixed foods and meals, and ability to quantify the extent of food processing among the large and otherwise homogeneously categorized groups of ultra-processed foods. Given that our algorithm only needs the Nutrition Facts, information already accessible to consumers on packaging and via smartphone apps, web portals, and grocery store and restaurant websites, *FPro* can help monitor the reliance of an individual’s diet on less or more processed food. Building on the portability of our model, we were able to extend the *FPro* assessment to over 50,000 products collected from major US grocery store websites, a first step towards a systematic characterization of the food environments [82].

Differently from dietary indexes such as HEI-15 [83], designed to measure the alignment of individuals’ diets with the 2015-2020 Dietary Guidelines for Americans, *iFPro* and *FPro* help us identify which foods to substitute, to shift individual consumption patterns towards a less processed diet. Our substitution heuristic indicates that minimal changes in diet can significantly reduce disease risk, a strategy hard to implement with the current NOVA classification, which classifies more than 70% of the food supply as NOVA 4 [84].

The consistent predictive power of *FPro* in the epidemiological analysis indicates that it offers an accurate global scale of food processing, capturing the chemical-physical alterations of food and its impact on health. However, *FPro* is currently best suited to rank foods within the same food category, hence offering accurate input for substitution strategies, as explored above. In other words, we should first identify a chemically-driven food group (e.g. “Fruit”), and then quantify the extent of nutrient alterations that leads to different degrees of processing.

Overall, a combination of *FPro* with epidemiological studies and food classification could lead to an automated and practical pipeline capable of systematically improving population diet and individual health.

## Data Availability

All codes and data necessary to train and test the algorithm are available at https://github.com/menicgiulia/MLFoodProcessing.
Additional codes are available upon request at. All datasets can be accessed through the cited references.

## Acknowledgments

We thank Dr. Euridice Martinez Steele at the University of Sao Paulo for providing NOVA manual classification for FNDDS databases. We thank Dr. Wei Wang at Harvard Medical School for consulting as statistical expert. This work was conducted with support from Harvard Catalyst | The Harvard Clinical and Translational Science Center (National Center for Advancing Translational Sciences, National Institutes of Health Award UL 1TR002541) and financial contributions from Harvard University and its affiliated academic healthcare centers. The content is solely the responsibility of the authors and does not necessarily represent the official views of Harvard Catalyst, Harvard University and its affiliated academic healthcare centers, or the National Institutes of Health. A.-L.B is partially supported by NIH grant 1P01HL132825, American Heart Association grant 151708, and ERC grant 810115-DYNASET.

## Code and Data Availability

All codes and data are available at https://github.com/menicgiulia/MLFoodProcessing.

## Author contributions

G.M. and A.-L.B. conceived the project. G.M. designed the study, performed data modeling, analytical calculations, data query and integration, and wrote the manuscript. B.R. performed data query, data integration, statistical analysis, and contributed to writing the manuscript. D.M. contributed to interpreting the results and writing the manuscript. A.-L.B. contributed to the conceptual design of the study and writing the manuscript.

## SUPPLEMENTARY INFORMATION

## 1 Training Dataset

### 1.1 Food Composition Databases

The food supply, representing the full inventory of all foods available for human consumption, along with their nutritional content, plays an important role in determining an individual’s nutrient exposure. Nutritional information is captured by food databases, collections of nutrient measurements for extensive samples of the food supply. Here we define as nutrients all chemicals catalogued by food databases, whether they refer to unique chemicals, like vitamin C, or aggregate measures, like total fat or total sugar. Additionally, all major nutrient databases include calories, measuring how much energy our body could get from eating or drinking the selected product. Which foods and which nutrients to report is strictly dependent on the database considered. For instance, USDA SR Legacy, the authoritative source of food composition data in the United States contains 7,793 food items with variable nutrient resolution, from a minimum of 8 nutrients, up to 138 (Figure S1) [2]. In comparison, USDA FNDDS, designed for the epidemiological analysis of dietary intake data collected by the National Health and Nutrition Examination Survey (NHANES), reports 65 to 102 nutrients for all foods, depending on the edition, containing no missing nutrient values (Figure S1) [3, 4].

The nutrient resolution available to consumers is significantly lower: the Food and Drug Administration (FDA) mandates the listing of 14 nutrients on the nutrition facts label, from saturated and trans fat, to sodium and vitamin C [1] An updated nutrition facts label was finalized in 2016, removing vitamin A and C, but listing added sugars, vitamin D, and potassium. However, the compliance deadline for certain food categories was extended to July 2021, and for the majority of the data describing branded products, we observed a significantly higher coverage of the nutrition facts prior to 2016.

**Figure SI. 1:**
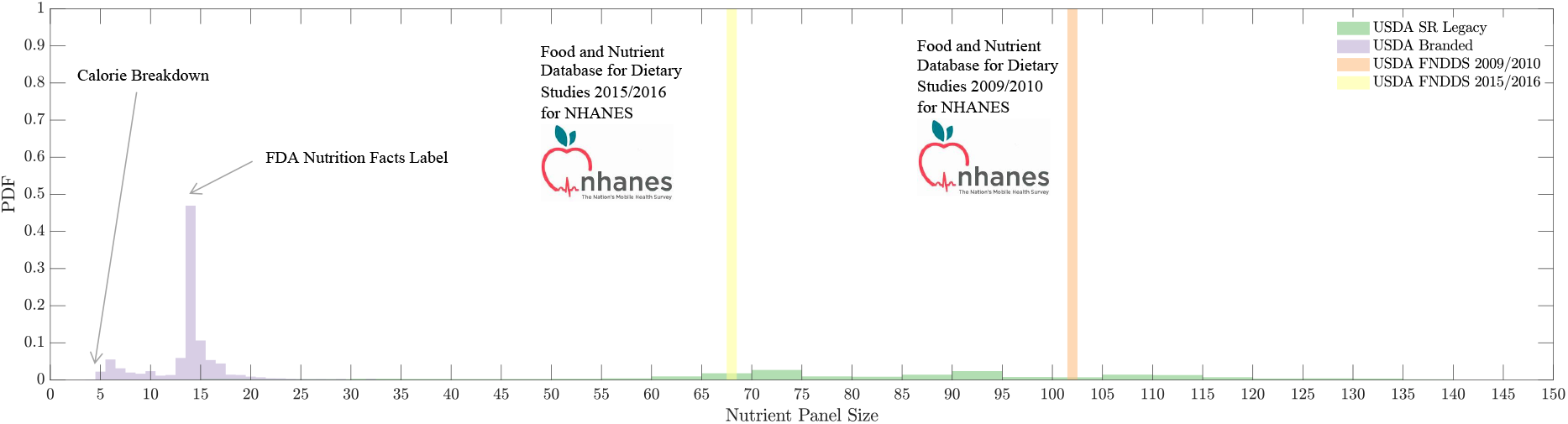
Nutrient Panel Resolution for Different Food Databases. To fully capture the nutrient alterations caused by food processing, we need access to the nutrient information characterizing each food in the food supply. The resolution available for branded products sold in grocery stores is very limited, frequently less than what required by FDA nutrition facts [1].

### 1.2 FNDDS 2009-2010

The Food and Nutrient Database for Dietary Studies (FNDDS) is designed by the USDA to provide food composition data (e.g. the amount of Vitamin C per 100 g of a selected ingredient) for foods and beverages reported in the dietary component of the National Health and Nutrition Examination Survey (NHANES), a biannual cross-sectional survey of the US Population conducted by Center for Disease Control and Prevention (CDC) to monitor the health of Americans. FNDDS is derived by combining the food items provided in the USDA National Nutrient Database for Standard Reference (SR). In other words, each item in FNDDS is related to one or more foods in SR, reported as ingredients in FNDDS. Differently from SR, designed for the dissemination of food composition data, FNDDS’s goal is to enable the analysis of dietary intake, hence it contains no missing nutrient values, an ideal setup to train machine learning models [5]. Since 2017, the USDA has been harmonizing these different data sources in FoodData Central (FDC), labeling SR data as SR-Legacy [6]. In particular, SR28 (released in 2015) is the final version of SR-Legacy databases, and it is the foundation of FNDDS 2015-2016. In addition to FNDDS and SR-Legacy, FoodData Central stores also Foundation Foods, a new food composition dataset that reports individual sample measurements behind the nutrient average values that populate the other databases, and metadata reporting the number of samples, location, time-stamps, analytical methods used, and if available, cultivar and production practices.

As shown in Figure S1, for the years 2007-2010 the USDA developed a flavonoids database for population surveys that extended the original nutritional panel of 65 nutrients to 102. For our analysis we kept all nutrients measured in g, mg or *μg*, dropping “Energy”, “Folate, DFE” and “Vitamin A, RAE”, resulting in 99 nutrients, converted to grams (g).

We chose FNDDS 2009-2010 as data training for FoodProX, as it gave us the possibility to combine the manual labels assigned by Steele et al. in [7], with the widest panel of nutrients available for population studies. Out of 7,253 foods in FNDDS 2009-2010, 2,484 food items are assigned to a unique NOVA class, while the remaining 4,769 foods are not classified (730), or need further decomposition (4,039) into 2,946 ingredients imported from the SR24 database.

Figure S2A shows the proportion of NOVA classes in the initial dataset, labels mainly derived by following the hierarchical encoding of food items provided by FNDDS. Indeed, each food is assigned to an 8-digit code, and the first five digits represent food categories. For instance, code 13230120 is assigned to “Pudding, flavors other than chocolate, ready-to-eat, sugar free” where the first digit ‘1’ represents “Milk and Milk Products”; the first two digits ‘13’ represents “Milk Desserts and Sauce”; and similarly ‘132’ represents “Puddings, Custards, and other Milk Desserts.” Relying on FNDDS food categories leads to two major limitations of the current classification system: (a) the existence of many exceptions. For example, a non ultra-processed food could belong to a category assumed to contain only ultra-processed food. Resolving these exceptions is a laborious work that requires domain knowledge; (b) limited scalability, as not all databases have a fine-tuned hierarchy of categories assigned to foods comparable to FNDDS.

Additionally, we found that some unclassified items, despite being labeled as requiring further decomposition, had only one ingredient. For instance, this is the case for the unclassified item ‘Egg, whole, raw’ (food code 31101010), created by linking only a single food from the SR database: ‘Egg, whole, raw, fresh’ (SR code 1123). Hence, we migrated 478 such unclassified single-ingredient foods to the training dataset (Figure S2B).

To further improve the training dataset, we manually classified nine foods, to extend the coverage of staple ingredients like ‘Salt’, or poorly represented classes like meat and fish (Table S1). In particular, the addition of ‘Salt’ in the training helped FoodProX to better calibrate the role of sodium in identifying ultra-processed food.

**Figure SI. 2:**
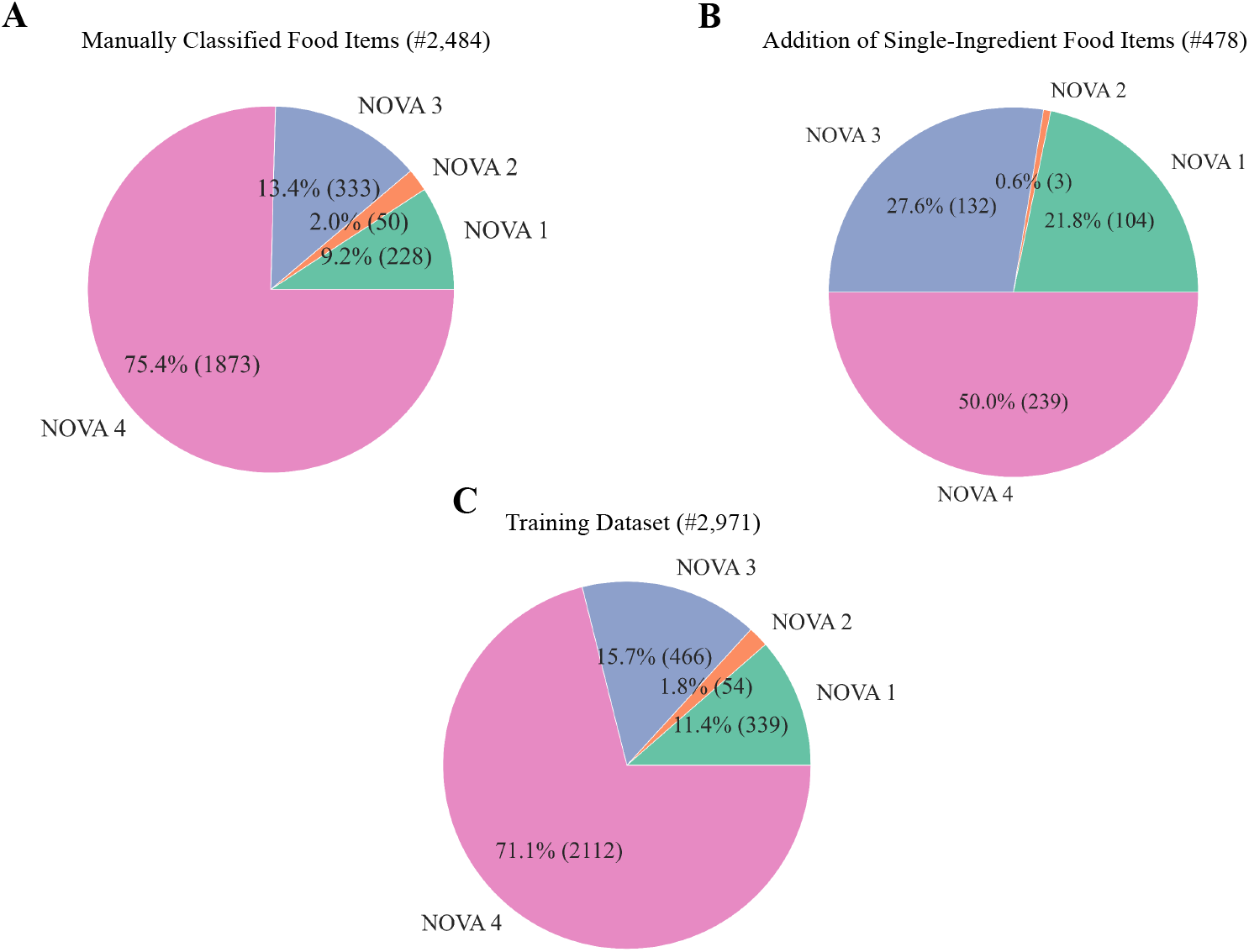
Proportion of NOVA Classes in the Manual Classification and Training Dataset. (**A**) Steele et al. [7] manually assigned NOVA classes to 2,484 from 7,253 food items in FNDDS 2009-2010, (**B**) We identified 478 single-ingredient and unclassified food items in FNDDS 2009-2010 linked to a single food in the SR database with assigned NOVA labels. (**C**) Final training dataset with the corrections on single-ingredient food items and the addition of 9 manually classified food items reported in Table S1.

**Table SI. 1:**
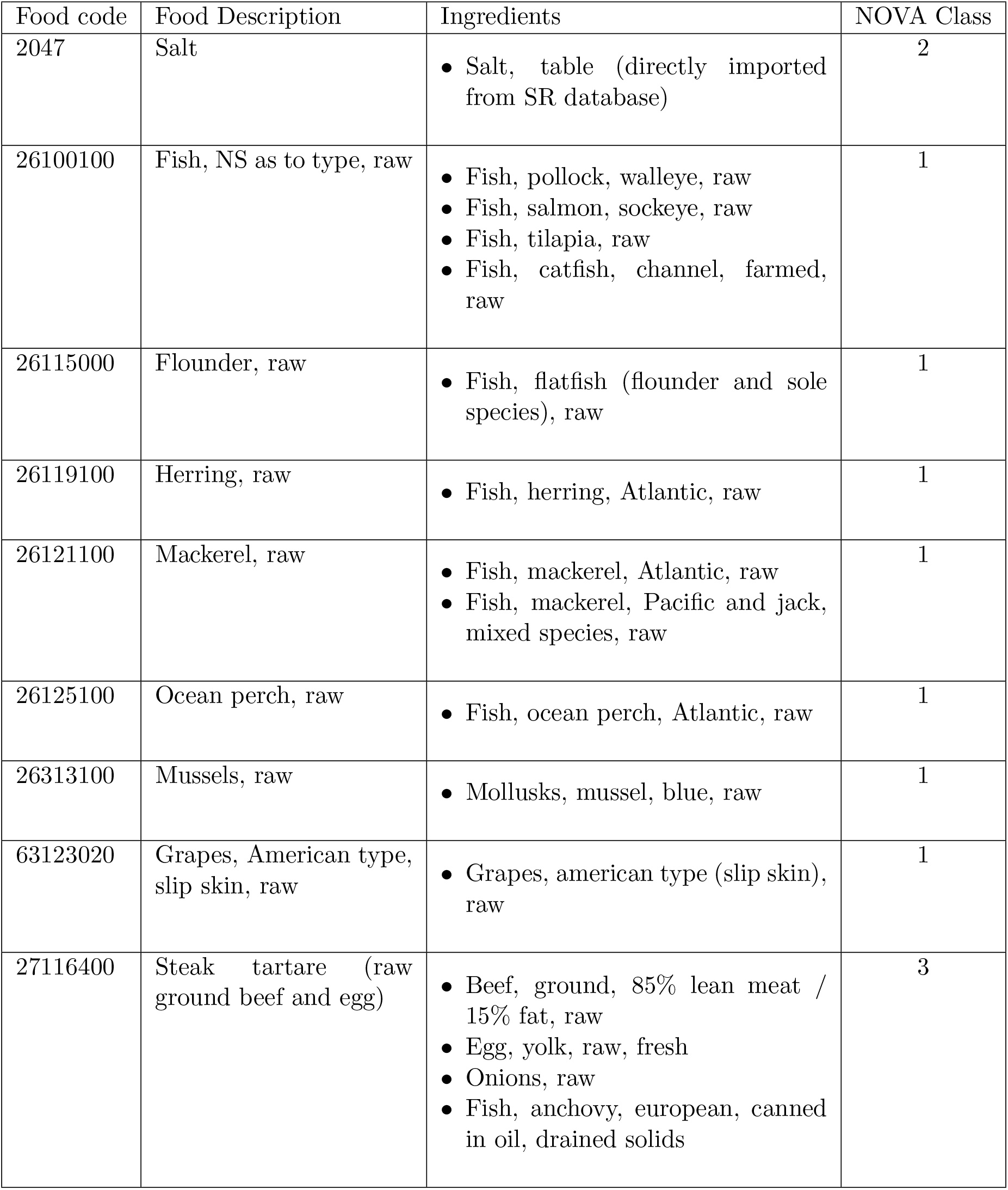
Manual Additions to the Training Dataset

### 1.3 NOVA Manual Classification Coverage

The coverage of NOVA classification for FNDDS 2001-2017 is presented by Figure S3. For over 55% of the databases, NOVA classification relies on having a precise ingredient decomposition of food items, an information that is extremely uncommon. To note, the manual NOVA classification has been updated since the initial classification on FNDDS 2009-2010 used in [7]. For clarity, only for FNDDS 2009-2010 in Figure S3, we used the NOVA classification conducted by Steele et al. in [7], which is the data-source that we used to train FoodProX. However, close to the convergence of this manuscript, Steele and colleagues have updated the manual classification for FNDDS 2009-2010, and propagated the labels in different cohorts by matching the food codes through the years. In the updated manual classification of FNDDS 2009-2010 the percentage of food items relying on ingredient decomposition is increased to 58.98% from 55.69%.

**Figure SI. 3:**
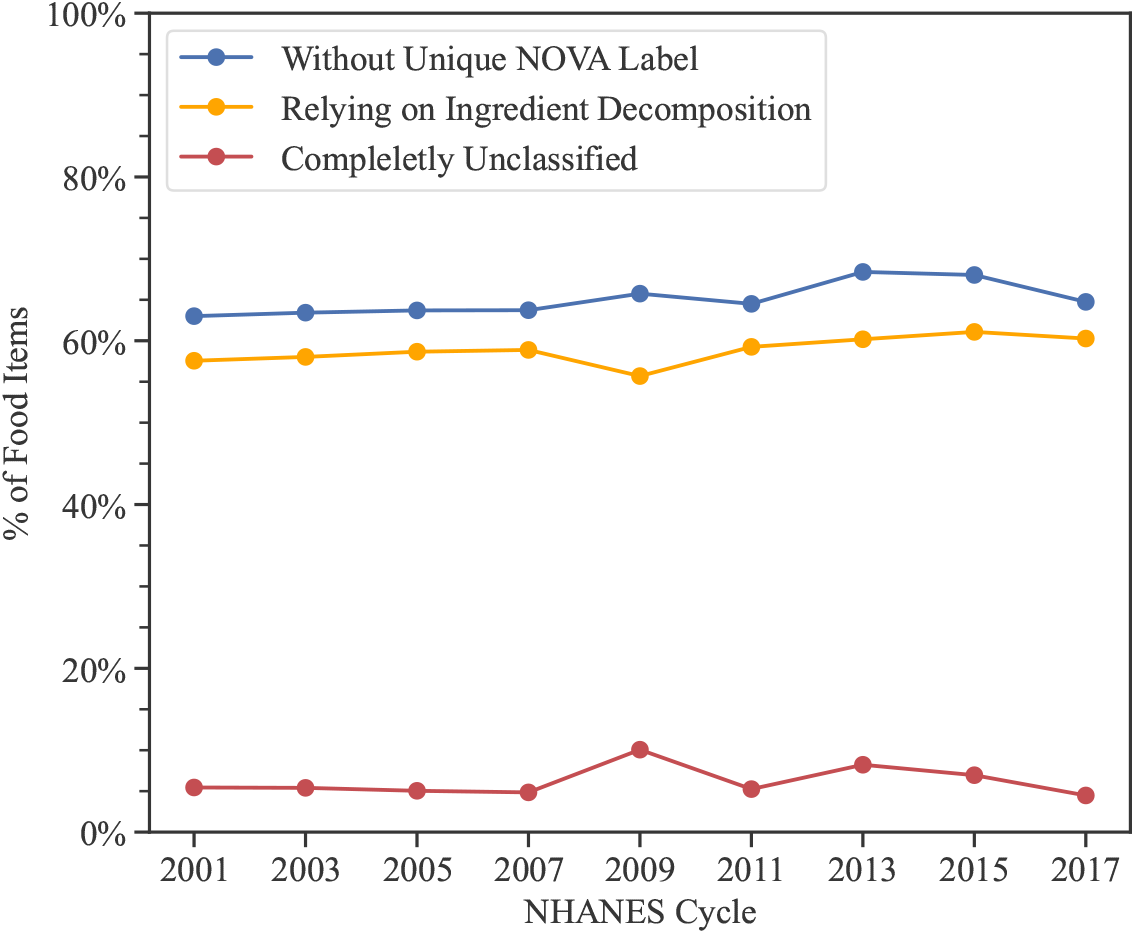
Manual NOVA Classification Coverage Over FNDDS 2001-2017. On average 35% of the food items have a manual NOVA label without relying on ingredient decomposition.

### 1.4 Nutrient Panels

The large nutritional panel available for FNDDS 2009-2010 allowed us to train FoodProX with varying subsets of nutrients. The widest panel consists of 99 nutrients, including the flavonoid measurements developed for NHANES 2007-2010 (Table S2) [8]. Among these 99 nutrients, we further selected and trained on 62 nutrients common to NHANES 2001-2018 (Table S3), and 58 nutrients available in NHANES 1999-2018 (Table S4). Figures 1A-1C in manuscript describe the results on FNDDS 2009-2010, with 99 nutrients, while Figures 1D-1E are related to the analysis of FNDDS 2015-2016, therefore using a nutrient panel of 62 nutrients. For the epidemiological analysis in Section 3, leveraging data from 1999 to 2006, we opted for 58 nutrients.

With the goal to tackle branded products and the consumer space, we additionally trained on a subset of 12 nutrients contributing to FDA nutrition facts (Table S5), excluding calories and total amount of trans fatty acids, as the latter is not available in the original batch of 99 nutrients.

All nutrients were log-transformed and 0 values were substituted with *e*^−20^, choice justified by the order of magnitudes spanned by nutrient concentrations in food [9].

**Table SI. 2:**
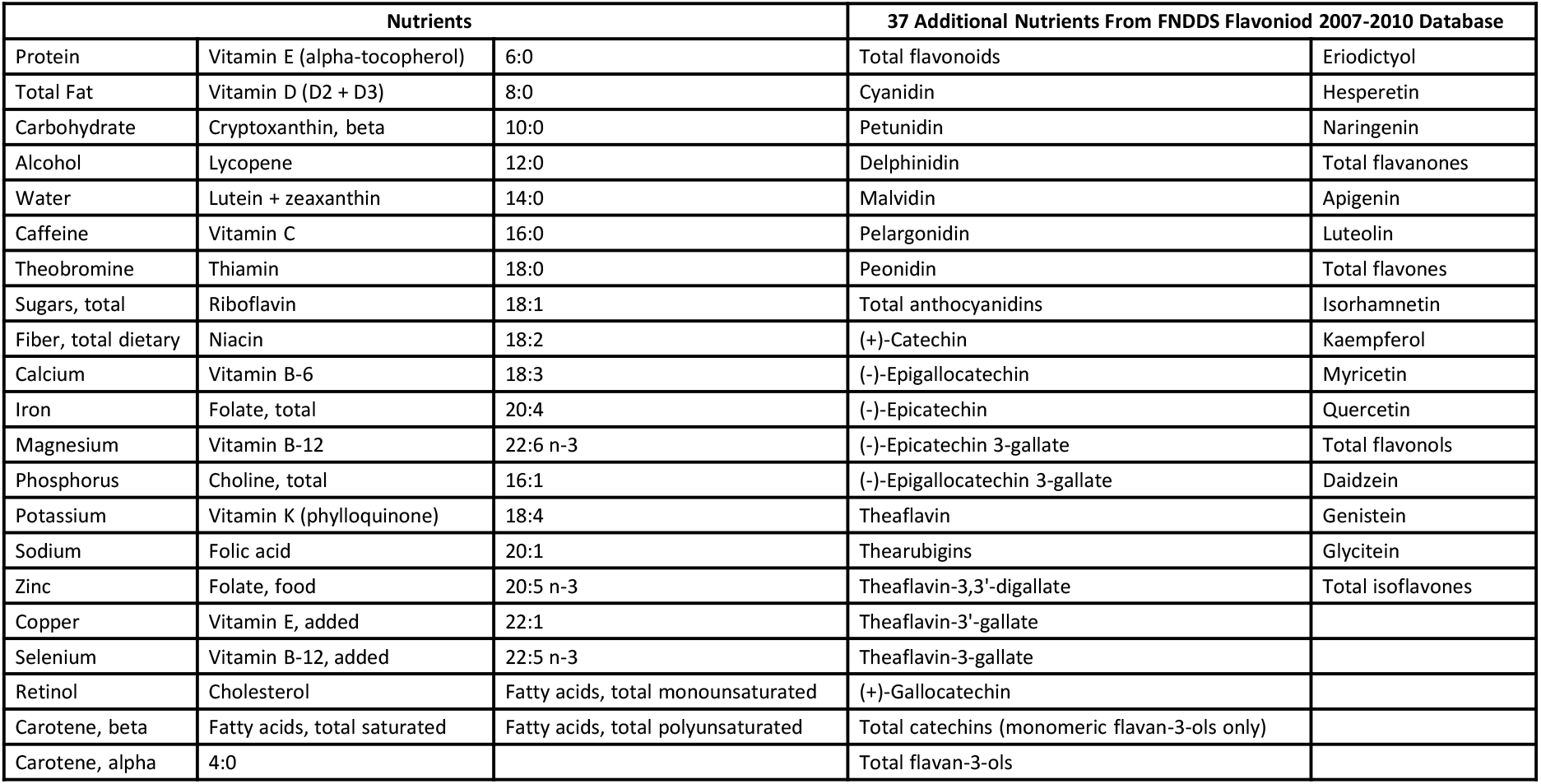
99 Nutrient Panel for NHANES 2007-2010

**Table SI. 3:** 62 Nutrient Panel for NHANES 2001-2018 Cycles

| Nutrients |  |  |
| --- | --- | --- |
| Protein | Vitamin E (alpha-tocopherol) | 6:0 |
| Total Fat | Vitamin D (D2 + D3) | 8:0 |
| Carbohydrate | Cryptoxanthin, beta | 10:0 |
| Alcohol | Lycopene | 12:0 |
| Water | Lutein + zeaxanthin | 14:0 |
| Caffeine | Vitamin C | 16:0 |
| Theobromine | Thiamin | 18:0 |
| Sugars, total | Riboflavin | 18:1 |
| Fiber, total dietary | Niacin | 18:2 |
| Calcium | Vitamin B-6 | 18:3 |
| Iron | Folate, total | 20:4 |
| Magnesium | Vitamin B-12 | 22:6 n-3 |
| Phosphorus | Choline, total | 16:1 |
| Potassium | Vitamin K (phylloquinone) | 18:4 |
| Sodium | Folic acid | 20:1 |
| Zinc | Folate, food | 20:5 n-3 |
| Copper | Vitamin E, added | 22:1 |
| Selenium | Vitamin B-12, added | 22:5 n-3 |
| Retinol | Cholesterol | Fatty acids, total monounsaturated |
| Carotene, beta | Fatty acids, total saturated | Fatty acids, total polyunsaturated |
| Carotene, alpha | 4:0 |  |

**Table SI. 4:**
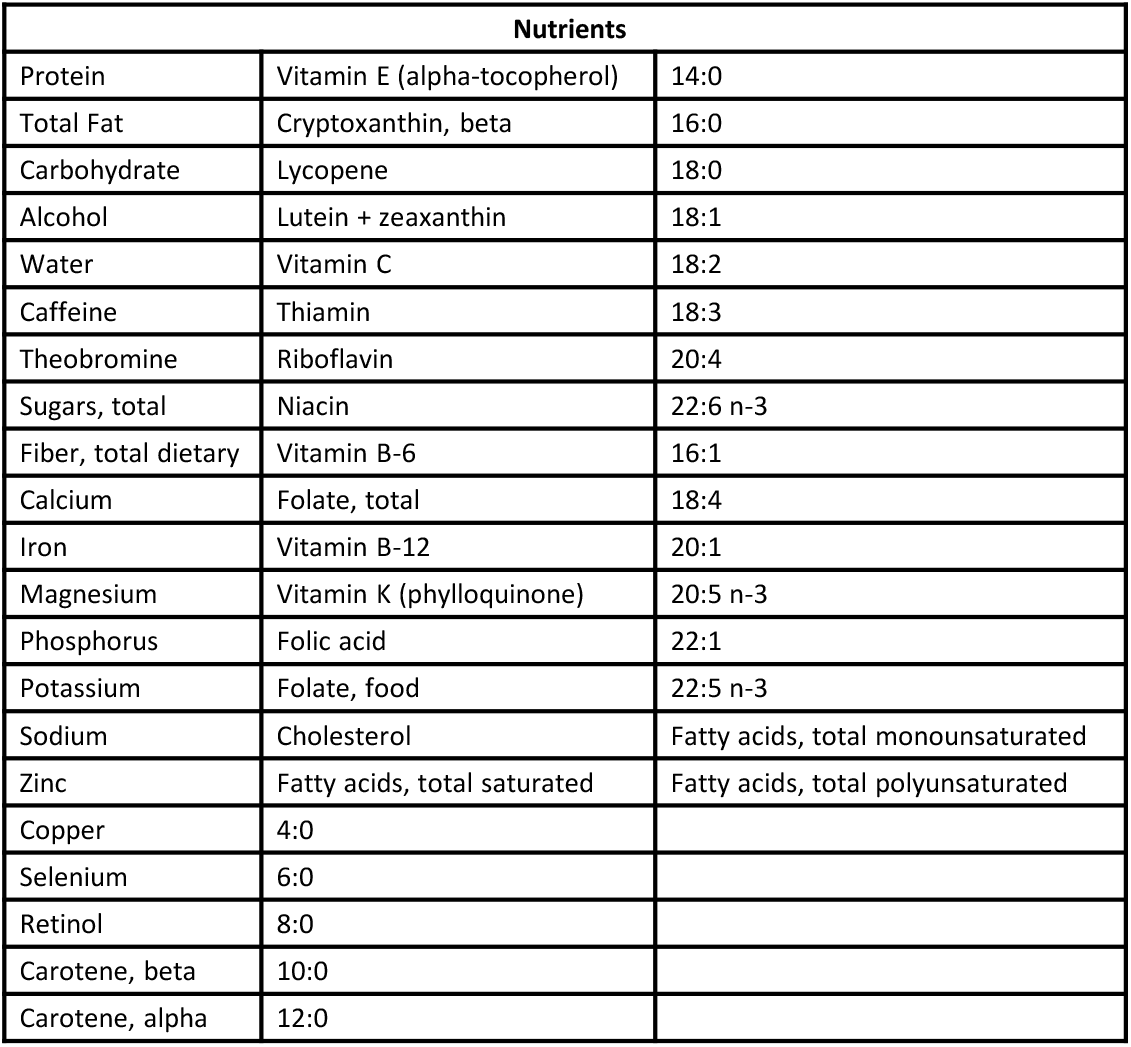
58 Nutrient Panel for NHANES 1999-2018 Cycles

**Table SI. 5:** 12 Nutrient Panel for Branded Products

| Nutrients |
| --- |
| Protein |
| Total Fat |
| Carbohydrate |
| Sugars, total |
| Fiber, total dietary |
| Calcium |
| Iron |
| Sodium |
| Vitamin C |
| Cholesterol |
| Fatty acids, total saturated |
| Total Vitamin A = Retinol + Carotene, beta + Carotene, alpha + Cryptoxanthin, beta |

### 1.5 Hierarchical Clustering of Foods according to Nutrients

Leveraging the 99 nutrient panel for FNDDS 2009-2010, we clustered all foods in an unsupervised fashion, using the dynamic tree cut algorithm [10]. The algorithm retrieves 20 clusters, annotated in the first column of the cluster-map shown in Figure S4. The nutrient-derived clusters are not good predictors of NOVA classes, both computationally-derived (Column 2) and manually-assigned (Column 3). We measure the mutual dependence between different clustering methodologies with adjusted mutual information (AMI), an adjustment of the classic mutual information (MI) to account for chance, that takes a value of 1 when two partitions are identical and 0 when the MI between two partitions equals the value expected by chance [11]. In particular, for the foods in the training data which were manually-assigned to NOVA 1, 2, 3, and 4, we find AMI=0.12. Similarly, for the classes predicted by FoodProX we obtain AMI=0.14. The nutrient-based hierarchical clustering is more consistent with the first two digits of the FNDDS food codes (Column 4), capturing broad food groups as defined by the database (AMI=0.39). This result suggests that the performance of FoodProX is not merely induced by the classification of foods based on nutrient content, but FoodProX combines in a non-linear fashion the features of processing techniques (supervised information learned from NOVA manual labels), with food composition data (unsupervised information learned from FNDDS nutritional values). Similar results were obtained with 62, 58, and 12 nutrient panels described in Section S1.4.

**Figure SI. 4:**
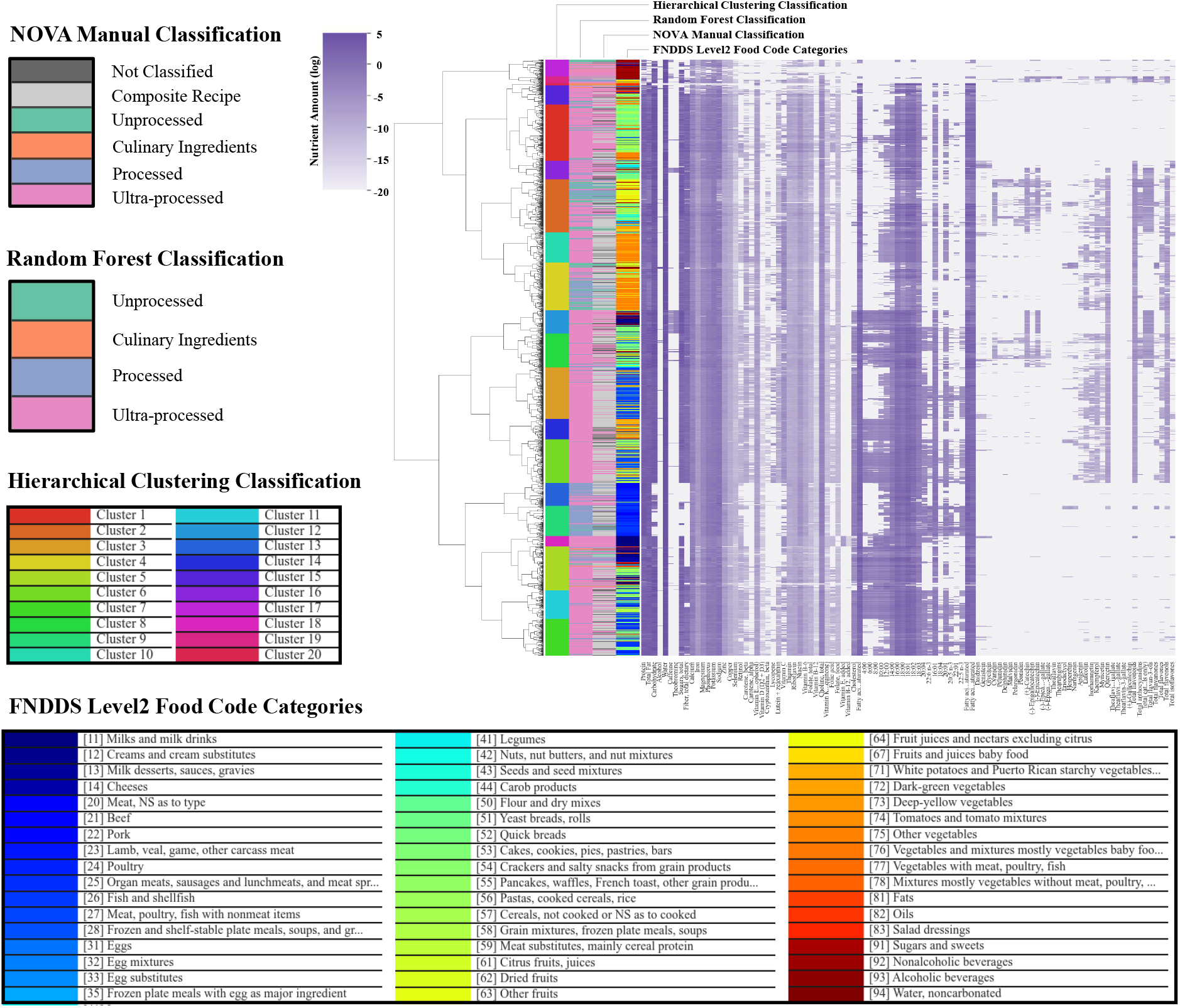
Hierarchical Clustering of Foods according to Nutrient Content. We clustered all foods in FNDDS 2009-2010, each represented by a vector of 99 log-transformed nutrients. The cluster-map is annotated according to different classification strategies. In Column 1, starting from the left, each color corresponds to a cluster found by the dynamic tree cut algorithm. In Column 2 we report the predicted NOVA classes by FoodProX, while Column 3 encodes the manual labels used during the training phase of FoodProX. Finally, in Column 4 items are color-coded according to the first two digits of their FNDDS food codes.

## 2 Random Forest Classifier FoodProX and Food Processing Score *FPro*

### 2.1 Random Forest Classifier

FoodProX is based on a Random Forest Classifier whose hyper-parameters were chosen using the python *sklearn RandomizedSearchCV* function with a 5-fold stratified cross validation. In particular, the sampled search considered a number of trees between 200 and 2000, a maximum number of features equal to √ or to log_2_, and a maximum depth of the trees between 100 and 500. Over 100 random samples, the function picked a number of estimators equal to 200, a maximum number of features equal to √, and a maximum tree depth equal to 420 (currently used). Further runs of the random search found other combinations of parameters spanning the whole search intervals, suggesting an overall robust performance of the classifier, independently from the hyper-parameter tuning.

We evaluated the performance and stability of FoodProX over a 5-fold stratified cross validation of the labeled dataset (Figure S13C), with varying input resolution. In Figures S13A-H we show the ROC curves and Precision-Recall curves for each NOVA class, while in Tables S6A and S6B we report average and standard deviation of AUC and AUP over the 5 folds, and across the different nutrient panels, as reported in the manuscript. The high performance for different nutrient resolutions is encouraging, as for many foods we lack access to an extensive panel of nutrients.

To improve the performance of the classifier on new data and limit over-fitting, we retrained it using SMOTE [12] to correct for the unbalance in class representation, and created and ensemble voting system of 5 classifiers trained on 4/5 of the generated data. The predictions on unseen data are then calculated as the average of the 5 classifiers.

**Table SI. 6:** AUC and AUP for the four NOVA classes. For each NOVA class we report average and standard deviation of AUC and AUP over the stratified 5-folds, for 12, 62, and 99 input nutrients. We summarize the results across nutrient panels of different resolution in bold.

| (A) |  |  |  |  | (B) |  |  |  |  |
| --- | --- | --- | --- | --- | --- | --- | --- | --- | --- |
|  | NOVA 1 | NOVA 2 | NOVA 3 | NOVA 4 |  | NOVA 1 | NOVA 2 | NOVA 3 | NOVA 4 |
| Average AUC Nutrition Facts | 0.981662 | 0.966348 | 0.967094 | 0.976772 | Average AUP Nutrition Facts | 0.891112 | 0.756997 | 0.864605 | 0.990558 |
| Std AUC Nutrition Facts | 0.003621 | 0.044898 | 0.010411 | 0.003432 | Std AUP Nutrition Facts | 0.035105 | 0.169225 | 0.040566 | 0.001414 |
| Average AUC 62 Nutrients | 0.980837 | 0.962878 | 0.970605 | 0.979866 | Average AUP 62 Nutrients | 0.891971 | 0.736245 | 0.873025 | 0.991702 |
| Std AUC 62 Nutrients | 0.00173 | 0.048586 | 0.00806 | 0.004876 | Std AUP 62 Nutrients | 0.024412 | 0.185744 | 0.03922 | 0.002178 |
| Average AUC 99 Nutrients | 0.978806 | 0.960406 | 0.971156 | 0.980085 | Average AUP 99 Nutrients | 0.881419 | 0.74707 | 0.879123 | 0.991783 |
| Std AUC 99 Nutrients | 0.002772 | 0.052151 | 0.007394 | 0.004237 | Std AUP 99 Nutrients | 0.024092 | 0.193387 | 0.032601 | 0.001954 |
| **Average AUC** | 0.980435 | 0.963211 | 0.969618 | 0.978908 | **Average AUP** | 0.888168 | 0.74677 | 0.872251 | 0.991348 |
| **Std AUC** | 0.0012 | 0.002437 | 0.001799 | 0.001513 | **Std AUP** | 0.004785 | 0.008475 | 0.005952 | 0.00056 |

**Figure SI. 5:**
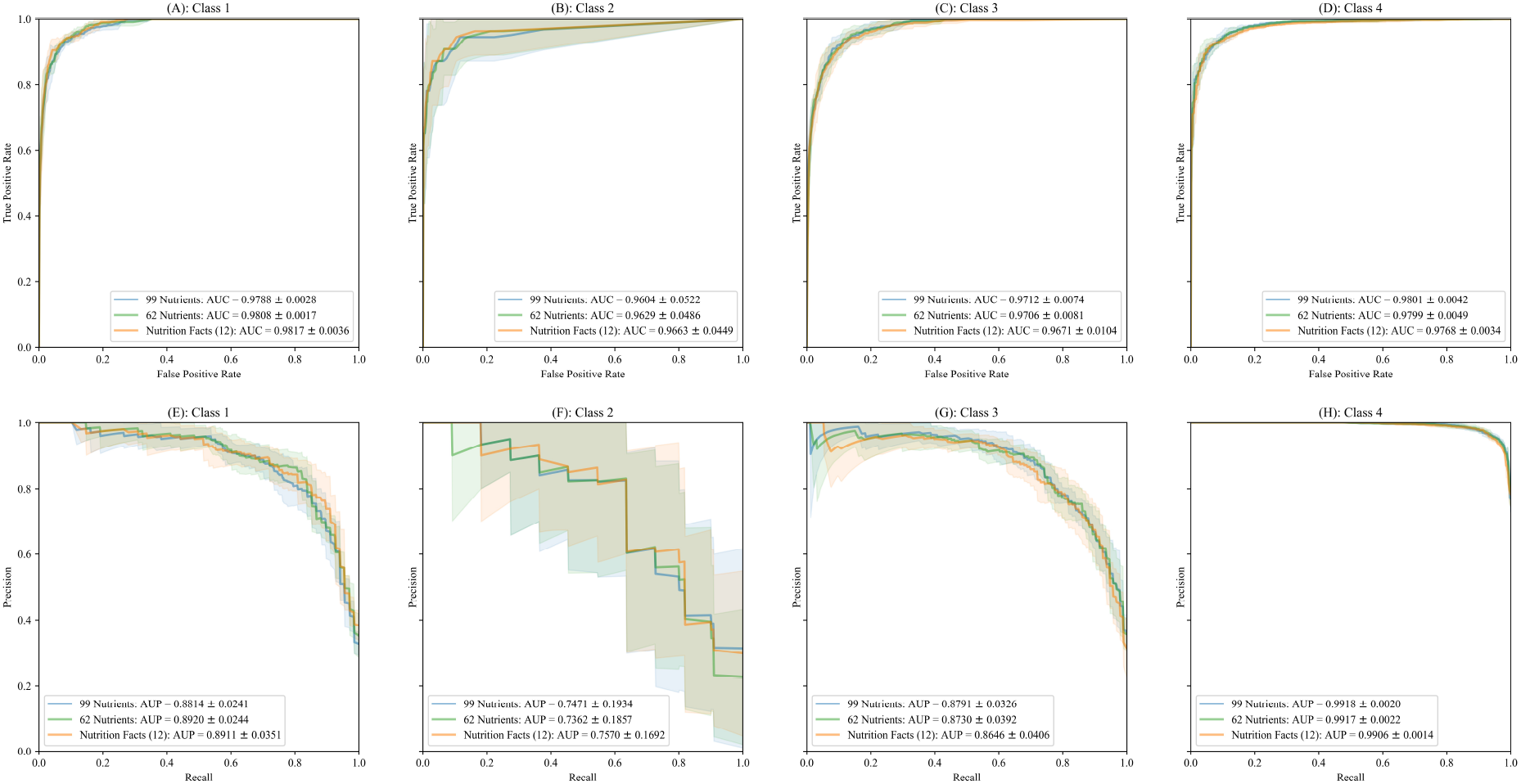
Random Forest Performance Over the 4 NOVA Classes. For each NOVA class we evaluated the performance of the random forest classifier with a 5-fold cross validation. We observe similar performances for the classifiers trained with 99, 62 and 12 nutrients. In Panels A-D we show the ROC curves for each NOVA class, while panels E-F display the Precision-Recall curves.

### 2.2 Feature Importance

Inspired by the work of Parr et al. [13], we investigated how different nutrients contribute to the overall performance of FoodProX, i.e. their feature importance. The most popular way to assess feature importance in the random forest algorithm is the mean decrease impurity, measuring how effective a feature is at reducing uncertainty (classifiers) or variance (regressors) when building the decision trees. However, this methodology is not reliable when potential predictors vary in their scale of measurement or their number of categories [14]. We opted for permutation feature importance, a technique quantifying the relevance of each feature by permuting the specific input column and measuring the decrease in accuracy or *R*^2^ compared to the baseline. This approach handles also the presence of collinear features, i.e. variables with some significant degree of linear or nonlinear dependence, that should be clustered and permuted together.

First, we modified the algorithm to work on a stratified 5-fold cross-validation, with data splits consistent with the cross-validation for the baseline model. Second, we addressed the high degree of collinearity of the nutrient space by removing all measurements like “Total Fat” or “Total flavonoids”, as they represent straightforward linear combinations of other nutrients. On the reduced set of 85 nutrients we studied both rank correlation and feature dependence, i.e. the extent in which each feature can be predicted by the others through a random forest regressor. We cluster together features with *ρ*_*Spearman*_ ≥ 0.80, and additionally, every nutrient well fitted by the random forest regressor is connected to each of the nutrients in the independent variable set that determines a drop ≥ 0.80 in the coefficient of determination (Figure S6A). Given the stochastic nature of the permutation feature importance, we repeated the estimation 20 times, and ranked the feature clusters according to their average drop in accuracy. In Figure S6B we report the top 10 most significant feature clusters, where only Sodium shows more predictive power than the other nutrients, suggesting that there is no single nutrient marker for food processing.

**Figure SI. 6:**
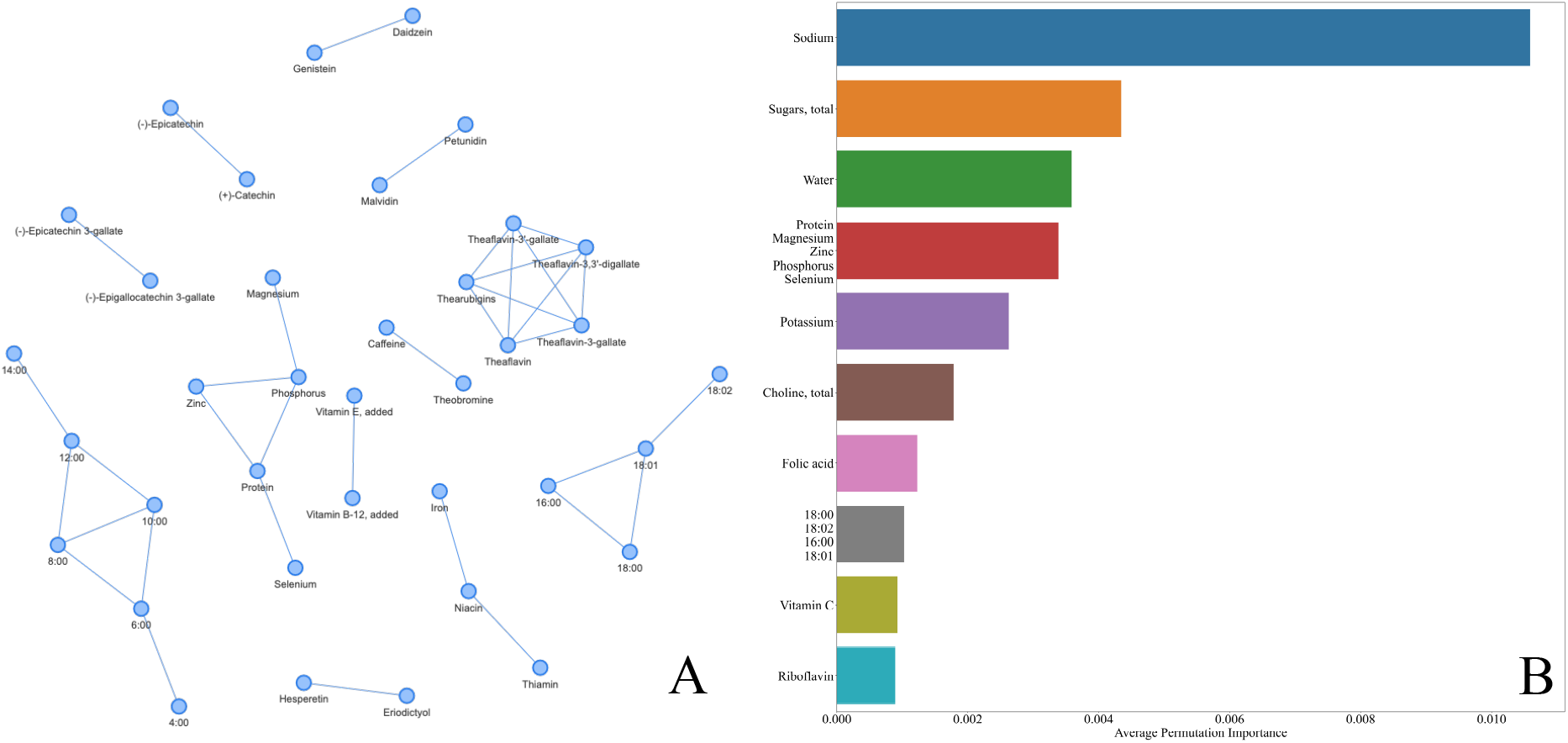
Permutation Feature Importance. (a) Clusters of features with strong dependence. We cluster together features with *ρ*_*Spearman*_ ≥ 0.80, and additionally, every nutrient well predicted by the remaining nutrient set through a random forest regressor is connected to each of the independent variables determining a drop≥ 0.80 in the coefficient of determination. Each connected component determines a single cluster of features permuted at the same time, while isolated nutrients (not shown) are permuted on their own. (b) Top 10 most important nutrient clusters sorted by average permutation importance over 20 reshuffles.

We further investigated the role of each nutrient *i* in determining the prediction for a selected food *f*, by using SHAP [15], over 5-fold stratified cross validation. The SHAP explanation method computes Shapley values from coalition game theory. The feature values of a data instance act as players in a coalition, and Shapley values indicate how to fairly distribute the “payout”, i.e. the prediction, among the features. For each food *f* and NOVA class *c*, SHAP specifies the explanation as

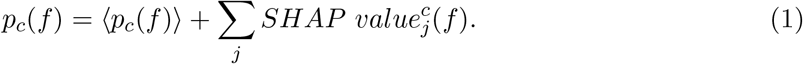

A player in the game could be also a group of features, as we previously investigated for the permutation feature importance analysis. However, due to high computational complexity, we focused on single nutrients, despite the presence of collinear features. In Figure S7 we show the top 10 nutrients in terms of 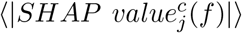 for each NOVA class *c*. The magnitude of the explanations varies significantly across NOVA classes, with only Sodium and Folic acid exhibiting distinct behaviors. For instance, when the amount of Folic acid is high, *p*_4_ is expected to be high as well, while the other NOVA classes display lower probabilities. In case of Sodium, high values are likely to increase *p*_4_ and decrese *p*_1_ and *p*_2_, while the processed class NOVA 3 displays mixed behaviors. Overall, we find that the ranking of SHAP values is in good overlap with Figure 6B.

To understand the statistical relevance of SHAP explanations, we introduced a new positive variable

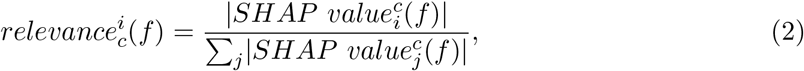

measuring to which extent nutrient *i* contributes to the superior limit of the absolute difference |*p*_*c*_(*f*)−*(p*_*c*_(*f*))|. In Figure S8, for each NOVA class *c* we display the violin-plot of 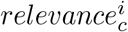 for the top 10 nutrients in terms of median effect. Each violin-plot is compared with the quantiles of a maximum entropy null model for feature relevance, i.e., a Dirichlet multivariate distribution with marginal probability for each feature equal to a beta distribution with parameters *α* = 1 and *β* = *n*_*features*_ − 1 [16]. The higher is the overlap between a violin plot and the null model ranges, the closer we are to a scenario lacking driving nutrients in determining the final class probability. In Figure S8, we observe a huge variability in the feature relevance of each nutrient, as captured by the shape of the violin-plots, evidence of the lack of strong driving signals in the model decision making. As expected, the largest nutrient contributions are found for NOVA 1 and NOVA 4, the two extreme processing classes.

**Figure SI. 7:**
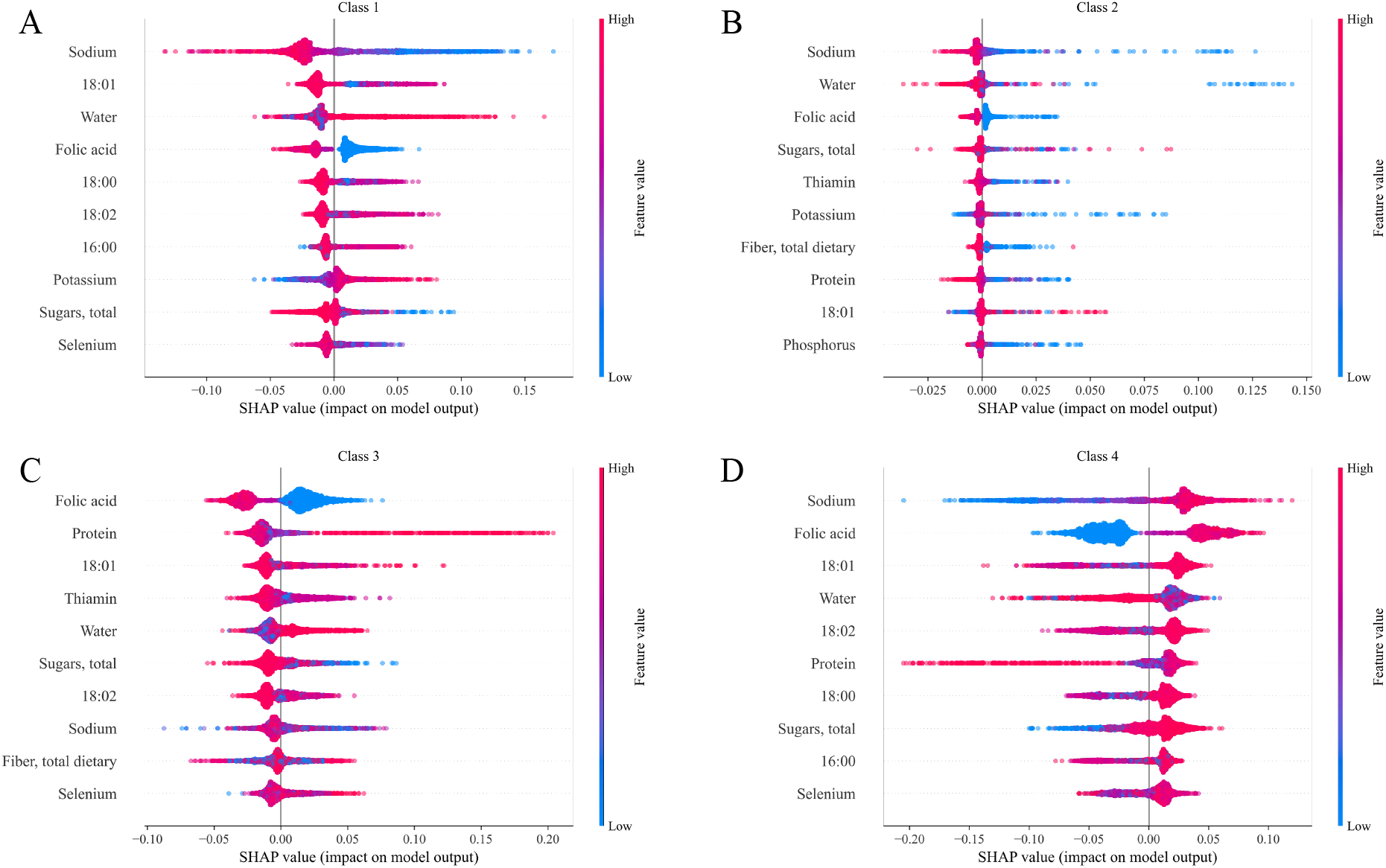
Shapley Values. Top 10 nutrients in terms of average 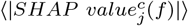 over all foods *f*, for class (a) NOVA 1, (b) NOVA 2, (c) NOVA 3, and (d) NOVA 4 (see Eq. S7). For each NOVA class *c*, the color scale correlates with the class probability *p*_*c*_.

### 2.3 Food Processing Score *FPro*

The classifier probability space is a 4-D probability simplex that collects all vectors satisfying

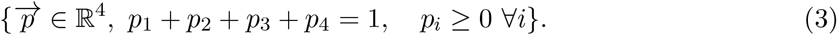

We define the processing score *FPro*_*k*_ as the projection of any food 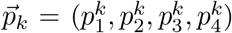 over the line going from the pure minimally-processed state 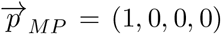 to the pure ultra-processed state 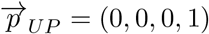, represented by the parametric equation

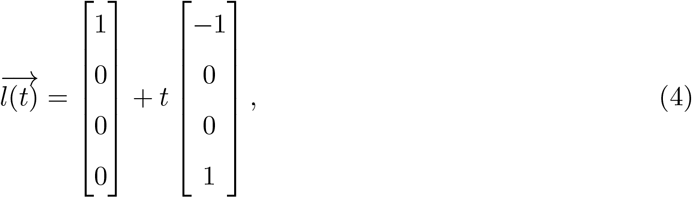

equivalent to the explicit equation *p*_1_ = 1 − *p*_4_. The orthogonal projection of food 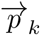 follows as the intersection between Eq. S4 and the plane passing through 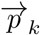 and orthogonal to 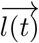, i.e.,

**Figure SI. 8:**
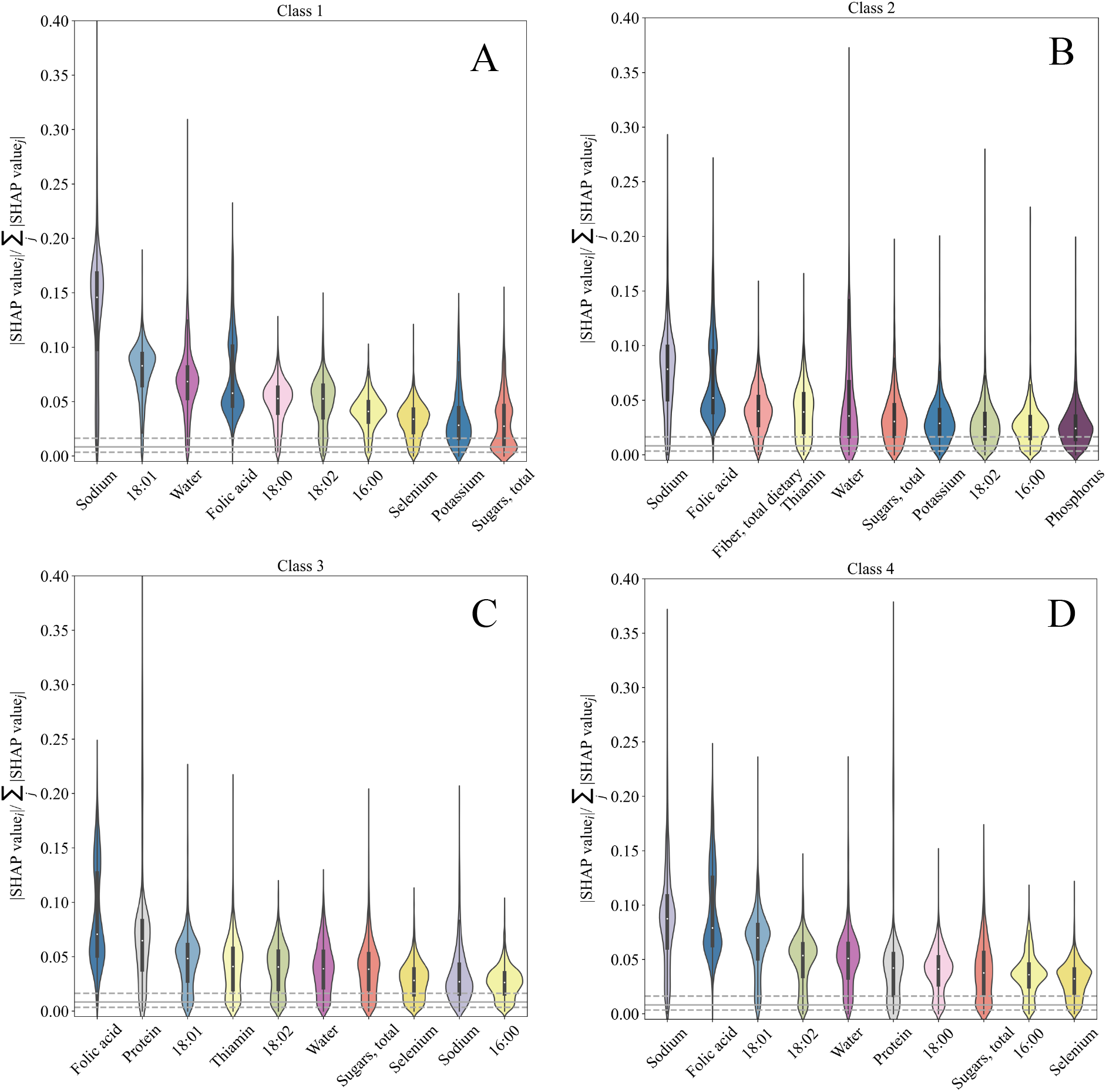
Relevance of Shapley Values. Top 10 nutrients in terms of median 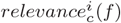 across all foods *f*, for class (a) NOVA 1, (b) NOVA 2, (c) NOVA 3, and (d) NOVA 4 (see Eq. S2). The grey dashed lines correspond to the quantiles *Q*_1_ and *Q*_3_ of a beta distribution with parameters *α* = 1 and *β* = *n*_*features*_ − 1. The full grey line points to the median of the same null model.

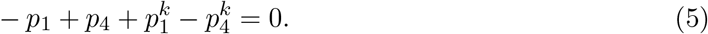

The parameter *t*^*^ satisfying Eqs. S4-S5 determines the processing score *FPro*_*k*_ in Eq. 1. Of note, *FPro* assigns a value around 0.5 for all NOVA 2/3 classes, which are then differentiated using the *p*_2_*/p*_3_ ratio.

We focused on the extreme classes NOVA 1 and NOVA 4, as by definition [17], they are the only ones with a clear “natural” ranking, ideal to define a processing scale. Indeed, NOVA 3 is not more processed than NOVA 2, they simply collect remarkably different items, according to nutrient composition and consumed portion. However, both classes are more complex than NOVA 1, and less processed than NOVA 4. While it is true that a “pure” NOVA 2/3 item would have *FPro* = 0.5, in real-world data, this is an uncommon scenario, so the extent of the residual probabilities *p*_1_ and *p*_4_ measures if a product is leaning towards the minimally processed or the ultra-processed extreme.

### 2.4 Validation of *FPro* in Different FNDDS Editions

We investigated the relation of *FPro*, trained on FNDDS 2009-2010 with a 62 nutrient panel, and NOVA manual classification in other editions of FNDDS, to control for any potential over-fitting and validate the performance of our algorithm on new foods, or foods whose nutrient content has changed over time. In particular, in Figure S9 we show the results for FNDDS 2015-2016, so far, the USDA database for dietary studies with the highest number of food items. All the four classes of manually labeled items correspond to well localized and distinguishable distributions of *FPro*.

**Figure SI. 9:**
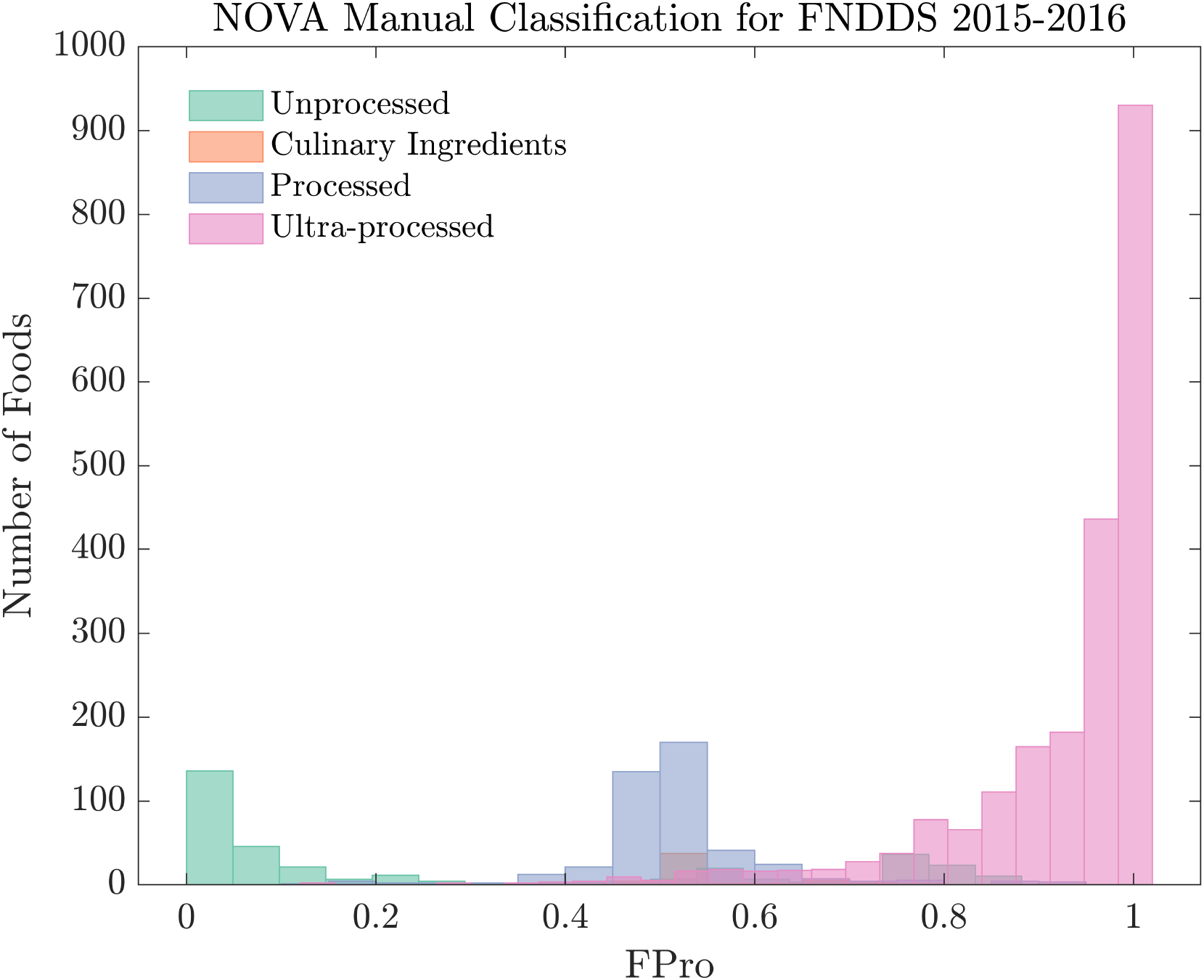
Food Processing Score for NOVA Manual Labels in 2015-2016. Variability of *FPro* (trained on FNDDS 2009-2010) within manual NOVA classes for FNDDS 2015-2016.

**Figure SI. 10:**
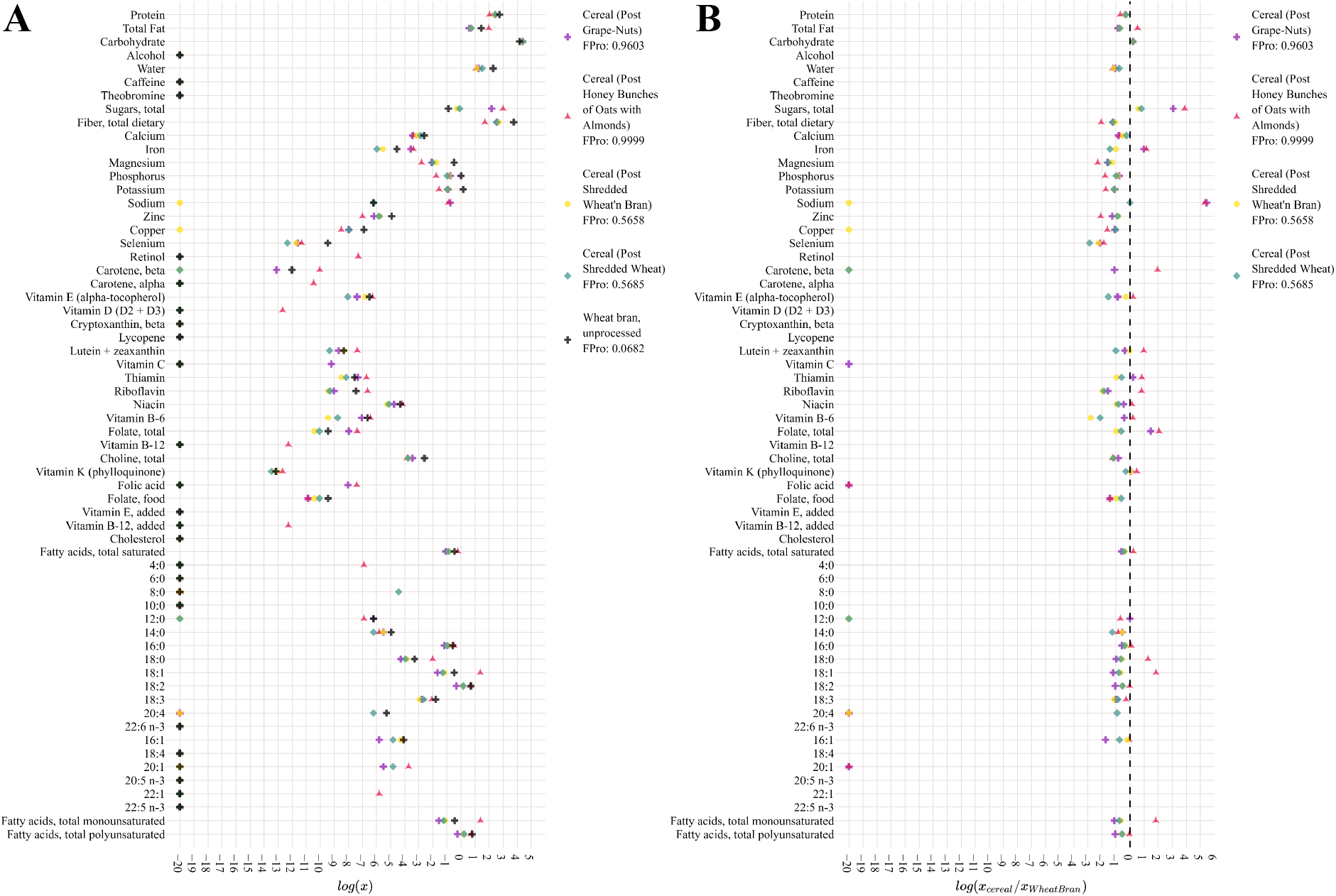
Nutrient Profiles of Post Cereals compared to “Wheat bran, unprocessed”. (a) All 62 nutrients measured in FNDDS 2015-2016 are shown in log-scale, and with varying color and marker depending on the food item. (b) We rescale the nutrient profile of each cereal by the corresponding value per 100 grams found in unprocessed wheat bran, and plot them in log-scale. The black dashed line corresponds to 1, i.e. identical nutrient content.

### 2.5 Case Study on Post Cereals

To further investigate the interplay between nutrient patterns and FPro, we collected the 62 nutrient profiles describing each Post cereal highlighted in Figure 2, and compared them with the nutritional values for 100 grams of unprocessed wheat bran (FPro=0.0682). We chose this specific ingredient as the Post Shredded Wheat ‘N Bran contains whole grain wheat, wheat bran, and the antioxidant Butylated hydroxytoluene [18], and we tried to find any potential matching item in FNDDS 2015-2016. In Figure S10, we compare the nutrient values in absolute terms (Panel A), or as ratio with their counterpart in wheat bran (Panel B). Interestingly, we observe how the pattern of alterations involves all nutrients, increasing the level of FPro even for simple products like Post Shredded Wheat ‘N Bran, as its nutrient profile is not characteristic of any natural ingredient per 100 grams, but it corresponds to a mildly processed food (FPro=0.5658).

### 2.6 Source of Food

The location where a food was prepared, as well the origin of the ingredients, could be indicative of its degree of processing. To investigate this hypothesis we used the variable DR1FS in NHANES, corresponding to the question “Where did you get (this/most of the ingredients for this)?”, and available for the years 2003-2018 (https://wwwn.cdc.gov/Nchs/Nhanes/2005-2006/DR1IFF_D.htm#DR1FS). By leveraging our analysis of the merged NHANES cohorts between 1999 and 2006 (Section S3.2), we were able to stratify FPro by food source for two cycles (2003-2004, 2005-2005). In Figure S11A we report the 25 different types of food sources found in the population, sorted by decreasing number of records. As expected, “Store” contributes to the majority of the records, followed by “Restaurant fast food/pizza”. In Figure S11B we selected 10 of the most popular food sources, and visualize the related distributions of FPro in increasing order of median. Overall, FPro is significantly different across source categories, as quantified by the Kruskal-Wallis H-test (p-value *<* 10^−15^), suggesting that the overlap between the source categories (driven by foods with multiple origins), while present, is limited. We also compared each pair of source categories with the Mann-Whitney U rank test, finding that (Cafeteria not at school, Bar/tavern/lounge), (Cafeteria at school, Bar/tavern/lounge), (Store, Bar/tavern/lounge), and (Restaurant with waiter/waitress, Bar/tavern/lounge) do not survive multiple testing with Bonferroni correction (*α* = 0.01), indicating continuous distributions with equal medians. Additionally, to control for “overpowering”, i.e. the scenario in which large samples almost surely determine a statistically significant outcome, we estimated the effect size *r* following [19]. Across the 45 combinations of food sources, 23 show *r* ≥ 0.1 (*small* effect), 6 have *r* ≥ 0.3 (*medium* effect), and the pair (“Grown or caught by you or someone you know”, “Vending machine”) is characterized by the largest effect size with *r* = 0.5798.

**Figure SI. 11:**
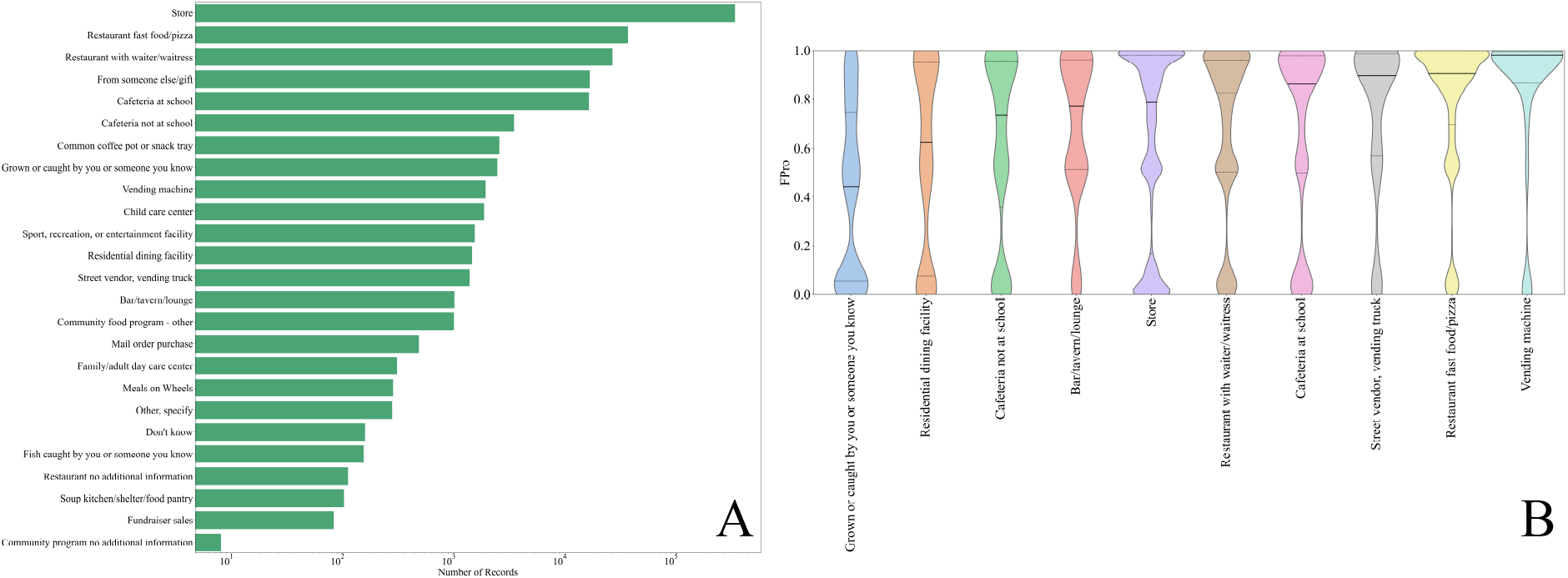
FPro stratified by food source in NHANES 2003-2005. (a) Food sources reported in the cohorts, sorted in descending order by number of records. (b) Violin plots of 10 selected food sources ranked in increasing order of median *FPro*. We annotate median values with full lines, and lower quartiles and upper quartiles with dashed lines.

## 3 Individual Diet Processing Scores *iFPro* and Exposome

### 3.1 Individual Processing Score *iFPro*

To capture the extent of processing in individuals’ diet we focus on two weighting schemes: a calorie-based score (Eq. 2), and a gram-based score,

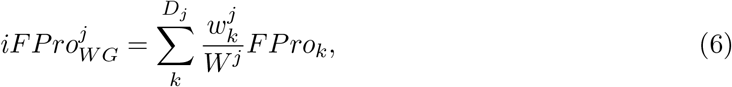

where *D*_*j*_ is the number of dishes consumed by individual *j, W*^*j*^ is her total amount of food in grams, and 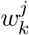 is the amount of grams consumed for each food item (excluding water consumption, see Section S3.4).

### 3.2 Population Characteristics

NHANES captures a variety of information ranging from demographic and dietary intake, to lab and physical examinations. This wealth of information is compiled into hundreds of publicly available data files, that altogether provide over 1,000 variables.

To investigate the relation between *iFPro* and health, we focused on NHANES 1999-2006 exposome and phenome database, a harmonized dataset created by Patel et al. in [20], merging 255 data files from four cycles of NHANES, for a total of 41,474 individuals and 1,191 variables. The summary statistics for *iFPro*_*W C*_ and *iFPro*_*W G*_, characterizing the 20,047 adults (18+) in the cohort, are presented in Tables S7 and S8. All predictions are calculated with the 58 nutrient panel in Table S4.

NHANES follows the well-established two-step 24HRs dietary recall interviews to sample the dietary intake of the American population [21, 22]. The first step is done in-person with a dietitian interviewing each applicant, while ensuring the highest quality of dietary recall over the past 24HRs. The second step consists of a phone interview within 3-10 days to capture a second dietary recall [23]. The multiple 24HRs dietary recalls have proved to be an effective method in the assessment of trends over the dietary intakes of individuals [24]. For all individuals who completed two-day dietary recalls (in-person and phone interview) we calculated a daily average *iFPro*, while for the remaining participants we used just the data from in-person interview. The relevance of each individual for population statistics is based on survey weights [25, 26].

**Table SI. 7:**
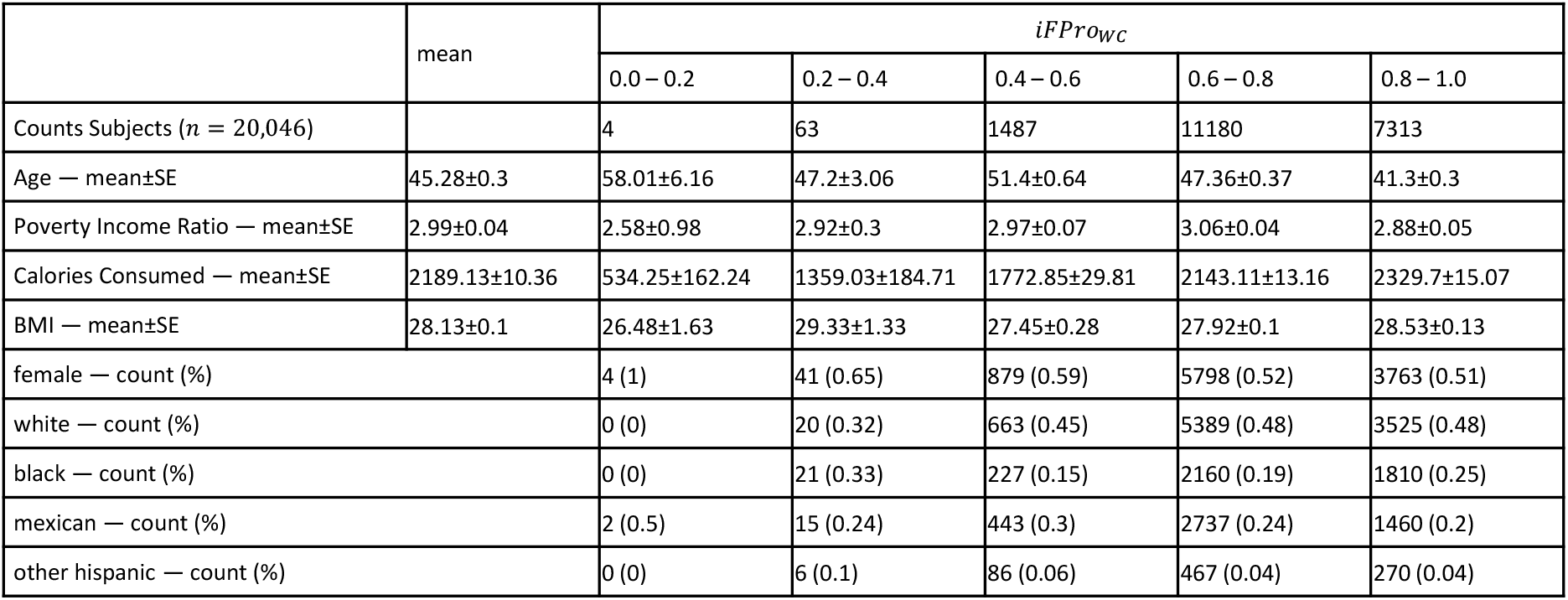
Population characteristics for. *iFPro*_*W C*_

**Table SI. 8:** Population characteristics for. *iFPro*_*W G*_

| | mean | $iFProWG$ | | | | |
| --- | --- | --- | --- | --- | --- | --- |
|  |  | 0.0 – 0.2 | 0.2 – 0.4 | 0.4 – 0.6 | 0.6 – 0.8 | 0.8 – 1.0 |
| Counts Subjects ($n = 20,046$) | | 182 | 2699 | 7286 | 7230 | 2650 |
| Age — mean±SE | 45.28±0.3 | 52.81±1.55 | 54.78±0.61 | 49.11±0.38 | 41.5±0.34 | 34.71±0.3 |
| Poverty Income Ratio — mean±SE | 2.99±0.04 | 3.01±0.15 | 3.13±0.05 | 3.15±0.05 | 2.92±0.04 | 2.56±0.07 |
| Calories Consumed — mean±SE | 2189.13±10.36 | 1490.34±70.36 | 1821.13±19.67 | 2134.69±14.34 | 2342.11±17.73 | 2356.4±30.02 |
| BMI — mean±SE | 28.13±0.1 | 28.08±0.82 | 27.47±0.15 | 27.76±0.13 | 28.37±0.1 | 29.14±0.19 |
| female — count (%) |  | 112 (0.62) | 1618 (0.6) | 3851 (0.53) | 3594 (0.5) | 1310 (0.49) |
| white — count (%) |  | 107 (0.59) | 1682 (0.62) | 3790 (0.52) | 2921 (0.4) | 1097 (0.41) |
| black — count (%) |  | 20 (0.11) | 294 (0.11) | 1189 (0.16) | 1860 (0.26) | 855 (0.32) |
| mexican — count (%) |  | 37 (0.2) | 529 (0.2) | 1731 (0.24) | 1873 (0.26) | 487 (0.18) |
| other hispanic — count (%) |  | 9 (0.05) | 100 (0.04) | 295 (0.04) | 320 (0.04) | 105 (0.04) |

### 3.3 Correlation between *iFPro*_*W G*_, *iFPro*_*W C*_, and HEI-15

While we find an overall agreement in the population ranking determined by *iFPro*_*W G*_ and *iFPro*_*W C*_ (*ρ*_*Spearman*_ = 0.7029, Figure S12A), the two measures show significantly different patterns in epidemiological associations, in particular regarding chemical exposures (Figure S20). Indeed, a weight-based index could capture complex dietary patterns arising from consumption of highly processed beverages such as zero-calorie soft drinks, or any type of food contaminants whose amount is independent from the provided calories.

We compared *iFPro*_*W G*_ and *iFPro*_*W C*_ with the HEI-2015, a score measuring the alignment of an individual’s diet with the national dietary guidelines, ranging from 0 (no alignment) to 100 (complete alignment) [27]. We followed the National Cancer Institute (NCI) to calculate HEI-15 for NHANES participants [28]. As expected, we observe negative correlations emerging (Figures S12B and C), with both *iFPro*_*W G*_ (*ρ*_*Spearman*_ = −0.4862), and *iFPro*_*W C*_ (*ρ*_*Spearman*_ = −0.5575).

**Figure SI. 12:**
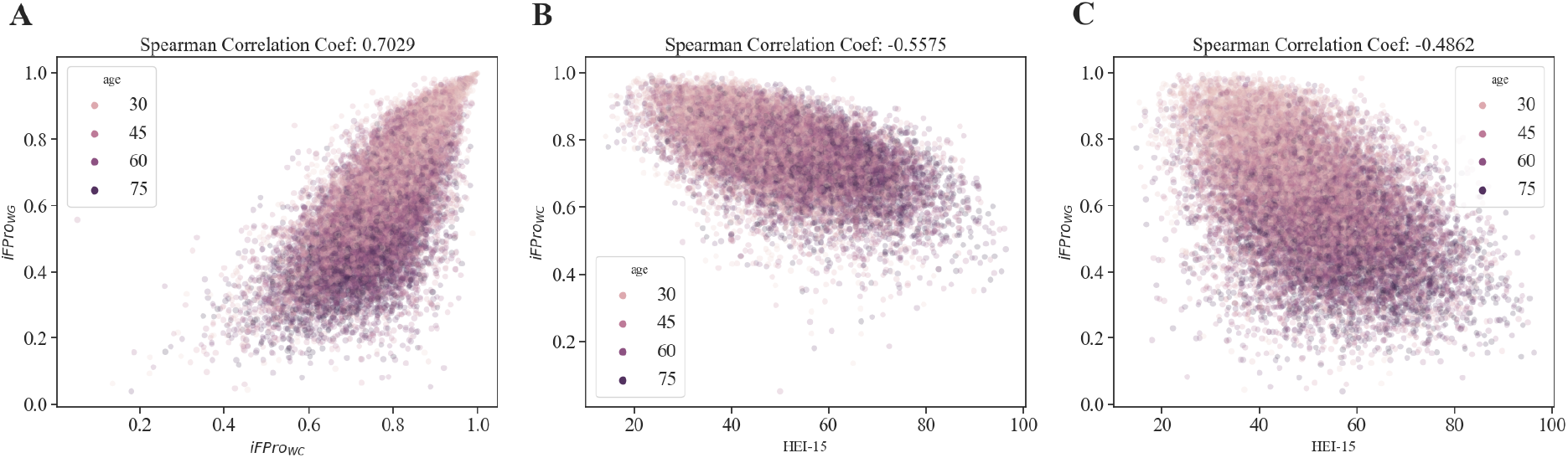
Spearman’s Correlation coefficients Between Two Variations of *iFPro* and Healthy Eating Index 2015.

**Figure SI. 13:**
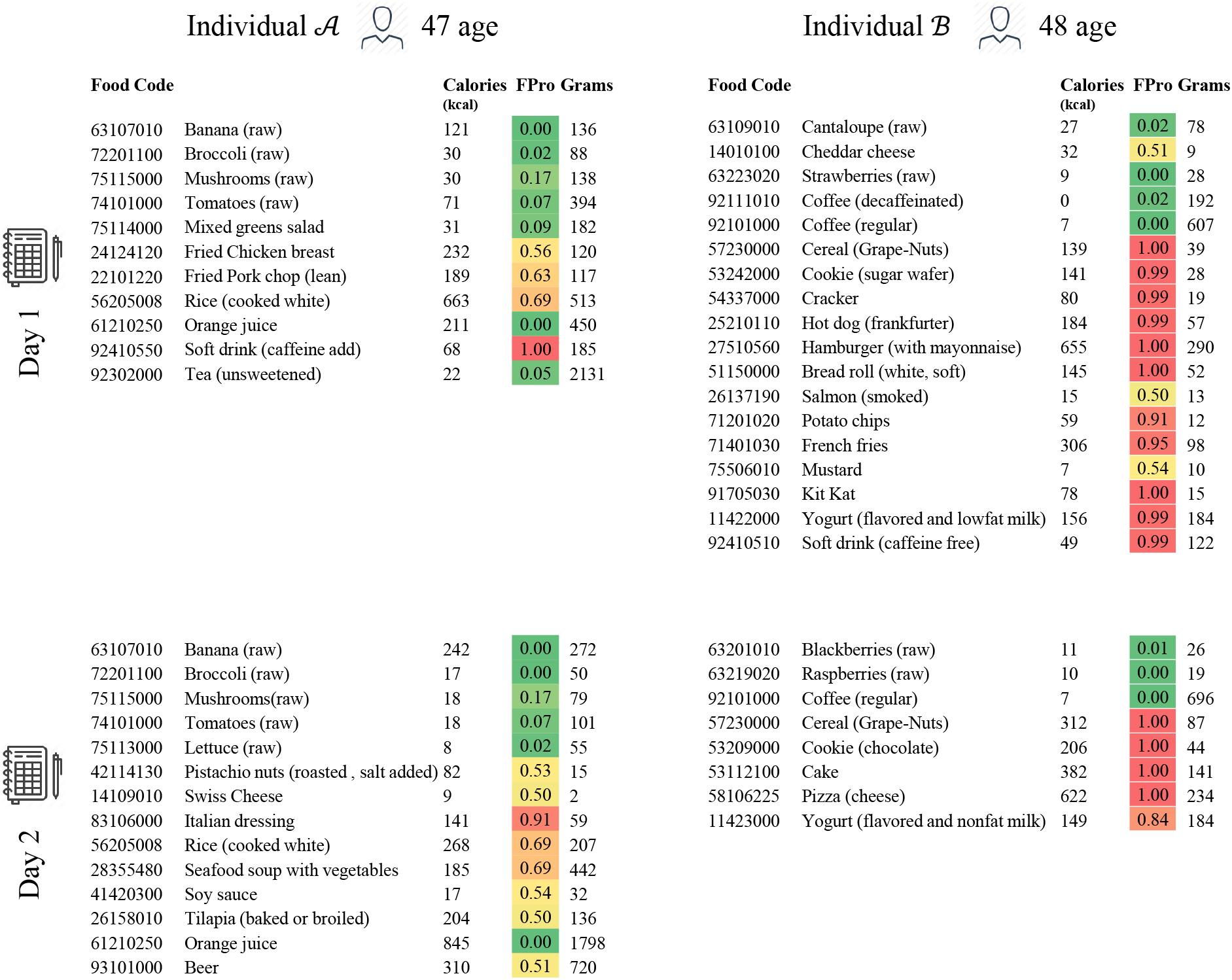
Dietary Recalls for Individual (SEQN 68484) and Individual (SEQN-ID 59440) annotated in Figure 2.

### 3.4 Water Consumption in NHANES

To calculate *iFPS*_*W G*_ (Equation S6), we removed four food codes regarding water consumption, as their reporting through different NHANES cycles showed inconsistencies. Indeed, we noticed that “Water as an ingredient” stopped being tracked since NHANES 2011-2012 (Figure S14 A), and the consumption of tap and bottled water started being recorded since NHANES 2003-2004 (Figures S14B and C). These inconsistencies would have affected our analysis of the pooled cohorts.

**Figure SI. 14:**
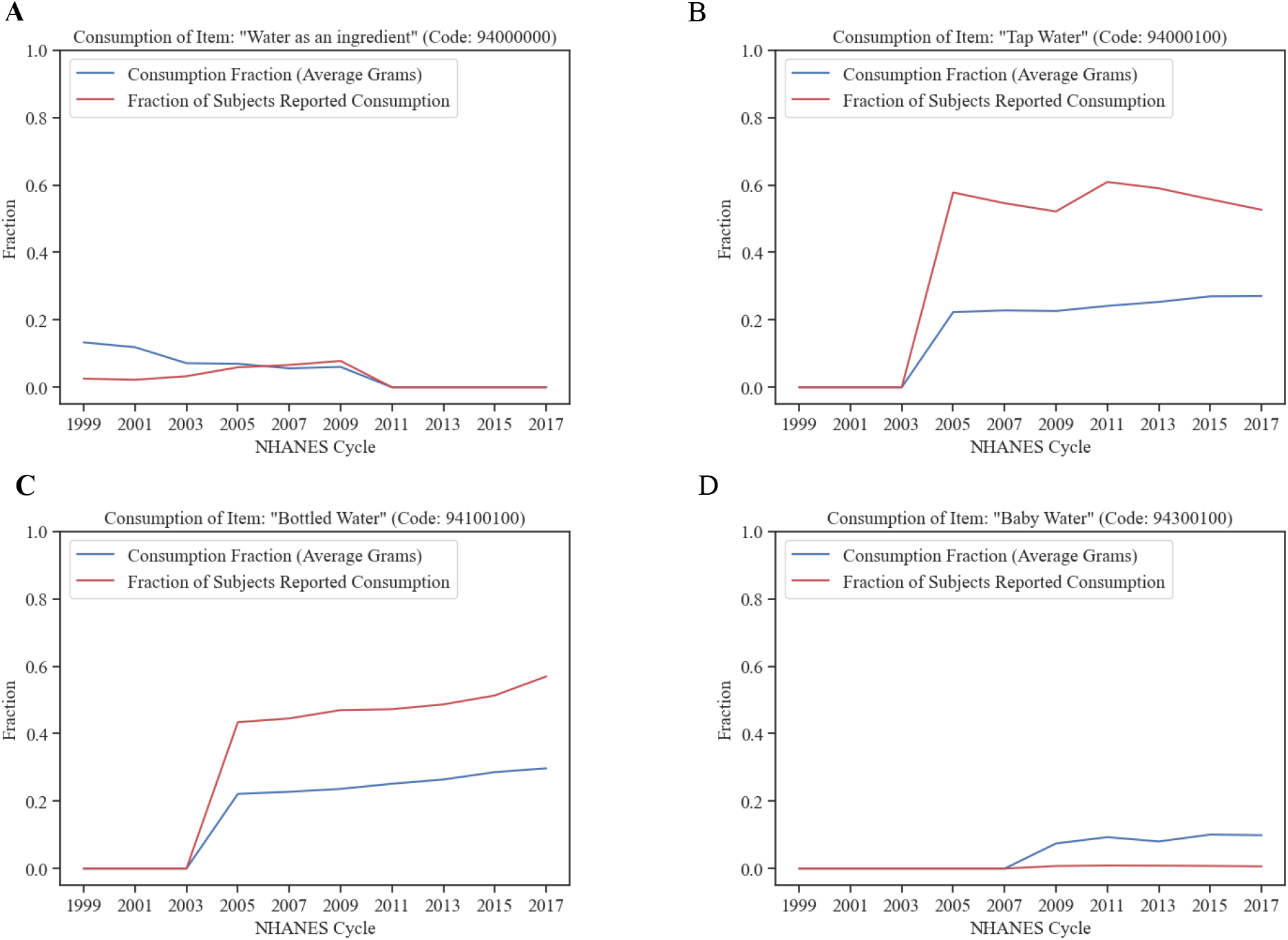
Water Consumption in Survey Data. (**a**) “Water as an ingredient” is no longer tracked since NHANES 2011-2012. (**b**-**c**) Bottled and tap water have been tracked since NHANES 2005-2006, hence this might introduce inconsistency when combining NHANES 1999-2004 cohorts with their succeeding cohorts. (**d**) The consumption of baby water has been tracked since NHANES 2009-2010.

### 3.5 Relation between *iFPro*_*W C*_ and WWEIA Food Categories

By leveraging our calculation of *iFPro*_*W C*_ (Eq. 2) over the merged NHANES cohorts between 1999 and 2006 (Section S3.2), we investigated the relation between trends in *iFPro*_*W C*_ and fraction of consumed calories in each of the What We Eat in America (WWEIA) food categories [29].

First, for each individual *j* we calculated the total fraction of calories contributed by food category *g*,

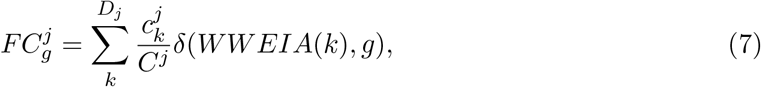

where *D*_*j*_ is the number of dishes consumed by individual *j, C*^*j*^ is the daily total amount of consumed calories, 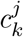 is the amount of calories contributed by each food item, and *δ* indicates the Kronecker delta, whose value is 1 when food *k* belongs to food category *g*, and otherwise 0. We proceeded in calculating the Spearman’s Rank Correlation between 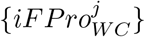 and 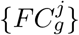 for each WWEIA class *g* across the cohort. Once accounted for multiple testing with Bonferroni correction (*α* = 0.01), we found a total of 64 WWEIA categories anti-correlated with *iFPro*_*W C*_, and 40 positively correlated (Figure S15).

We further investigated the representation of WWEIA categories in the first quintile *Q*_1_ of *iFPro*_*W C*_ (*iFPro*_*W C*_ ≤ 0.6908), compared to the last quintile *Q*_5_ (*iFPro*_*W C*_ ≥ 0.8585). These two sub-populations capture 4,010 individuals each, representing the most divergent dietary patterns in terms of ultra-processed food. The caloric consumption of each WWEIA category across the two sub-groups was tested with the Mann-Whitney U rank test, finding a total of 103 categories significantly changing from *Q*_1_ to *Q*_5_, once corrected for multiple testing (Bonferroni method with *α* = 0.01). To control for “overpowering”, as explained in Section S2.6, we calculated the effect size *r* following [19], and ranked all significant WWEIA categories accordingly (Figure S16A). Overall, the consumption of 21 food groups change between *Q*_1_ and *Q*_5_ with effect size ≥ 0.1, the baseline for small effect sizes. Among the biggest effect sizes we find “Soft drinks” (*r* = 0.4660), with average caloric fraction 5.61 bigger in *Q*_5_ compared to *Q*_1_, and “Bananas” (*r* = 0.2728), with average caloric fraction 14.07 bigger in *Q*_1_ compared to *Q*_5_ (Figure S16B).

**Figure SI. 15:**
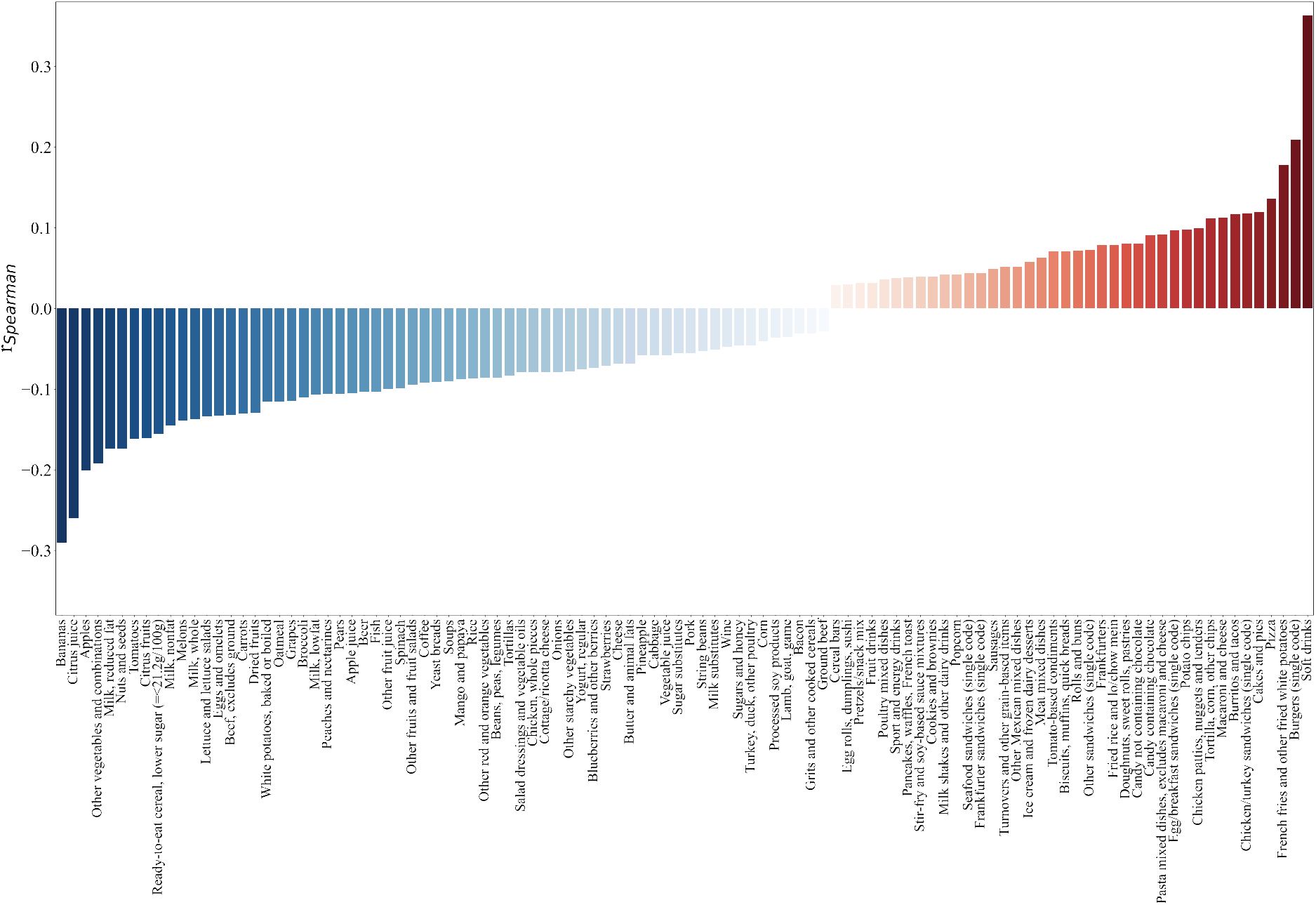
Spearman’s Rank Correlation between *iFPro*_*W C*_ and the caloric fraction contributed by each WWEIA category across NHANES individuals. 64 WWEIA categories significantly anti-correlate with *iFPro*_*W C*_ (blue), while 40 are positively correlated (red). The height of each bar is proportional to the correlation value, and all coefficients are sorted from the strongest anti-correlation to the strongest correlation.

## 4 Environment-Wide Association Study

Inspired by [30], we performed an Environment-Wide Association Study (EWAS) on the merged NHANES 1999-2006 cohort, to identify environmental factors and disease-related phenotypes associated with *iFPro*_*W C*_, *iFPro*_*W G*_, and fraction of calories contributed by manual NOVA 4. To do so, we collected data for 45 exposure modules in [20], and we further added one variable predicting diabetes according to fasting glucose levels ≥126 mg/dL, as advised by the American Diabetes Association [31], two variables predicting metabolic syndrome [32], two assessments of the Framingham Risk Score [33, 34], and the ACC/AHA Risk Score [35], quantifying the 10-year risk of non-fatal myocardial infarction (MI), congestive heart disease (CHD) death, or fatal or nonfatal stroke.

The variables are broadly categorized in two panels (Figure S17A): a health panel, gathering variables describing the overall health of the individuals, from biological age and nutrient biomarkers, to disease phenotypes, and a chemical panel, where we group all chemical exposures measured in blood or urines, linked to pesticides, contaminants, and processing chemical byproducts.

**Figure SI. 16:**
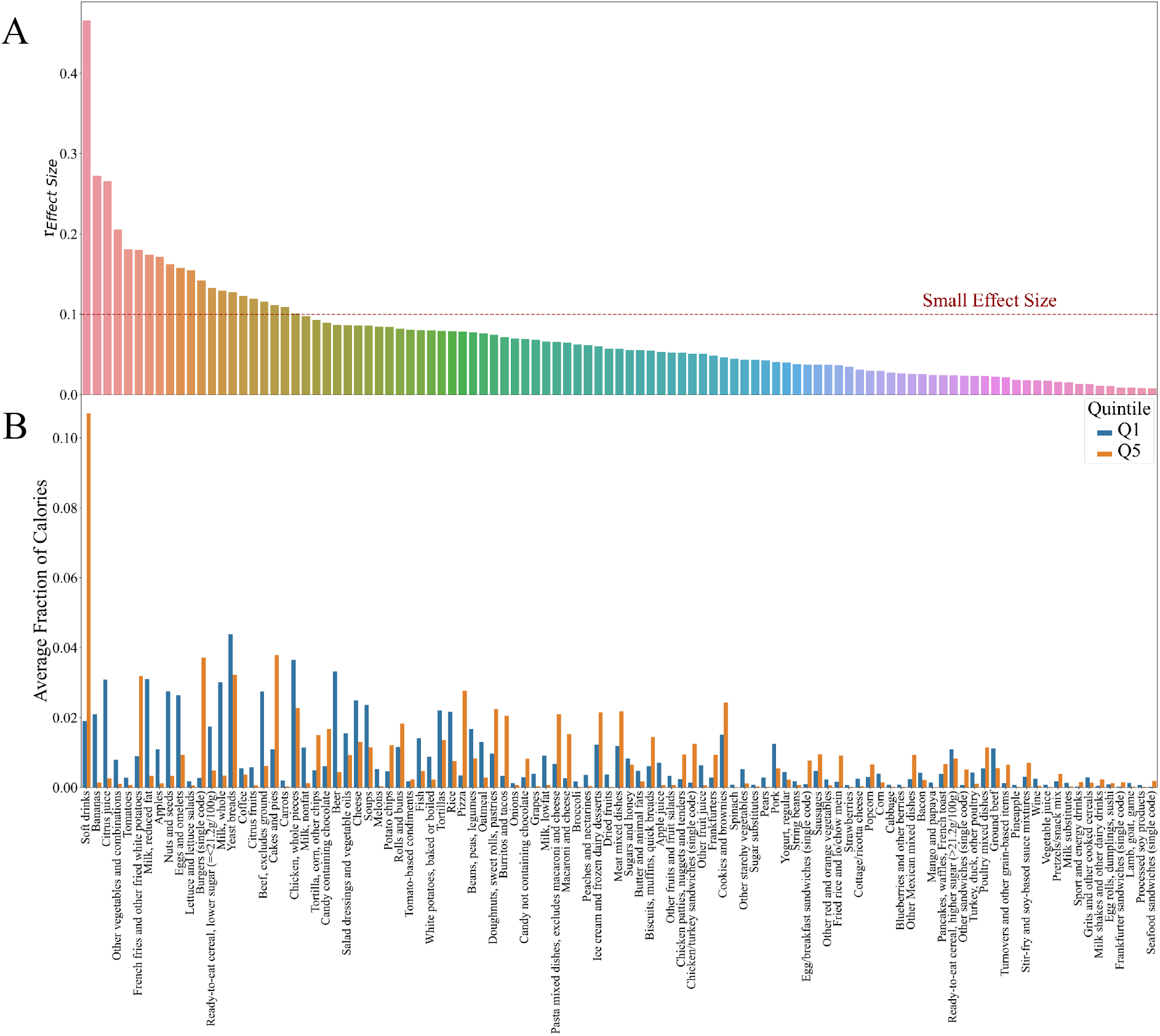
Comparison of *iFPro*_*W C*_ Extreme Quintiles *Q*_1_ **and** *Q*_5_. For individuals in the first quintile *Q*_1_ and in the last quintile *Q*_5_ of *iFPro*_*W C*_ we compare the caloric fraction contributed by each WWEIA category. In (a) we estimate the effect size *r* following [19], where *r* ≈ 0.1 is considered a *small* effect, *r* ≈ 0.3 a *medium* effect, and *r* ≈ 0.5 a *large* effect. The WWEIA food categories surviving multiple testing with Bonferroni correction (*α* = 0.01) are shown in decreasing order of effect size. In (b), following the same order, we show the average fraction of calories in *Q*_1_ and *Q*_5_.

To select the most robust signal, we studied only variables measured in at least two cycles of NHANES. To quantify the statistical associations, we employed the Survey-weighted Generalized Linear Model, and in particular, linear regression to predict continuous variables, and logistic regression for categorical (Figure S17B). All models were adjusted for age, sex, ethnicity, Body Mass Index (BMI), total-caloric intake, and estimated Socioeconomic Status (SES), as provided by NHANES. We employed the ‘survey’ statistical package in R to account for the complex survey design of NHANES [26]. We further filtered all categorical and continuous variables lacking a minimum sample size to perform regression analysis. In particular, for continuous variables we considered a ratio between number of covariates and number of data points ≤ 1/50, while for categorical we applied a similar threshold for the ratio between number of covariates and number of data points in the smallest category (Figure S17C).

**Figure SI. 17:**
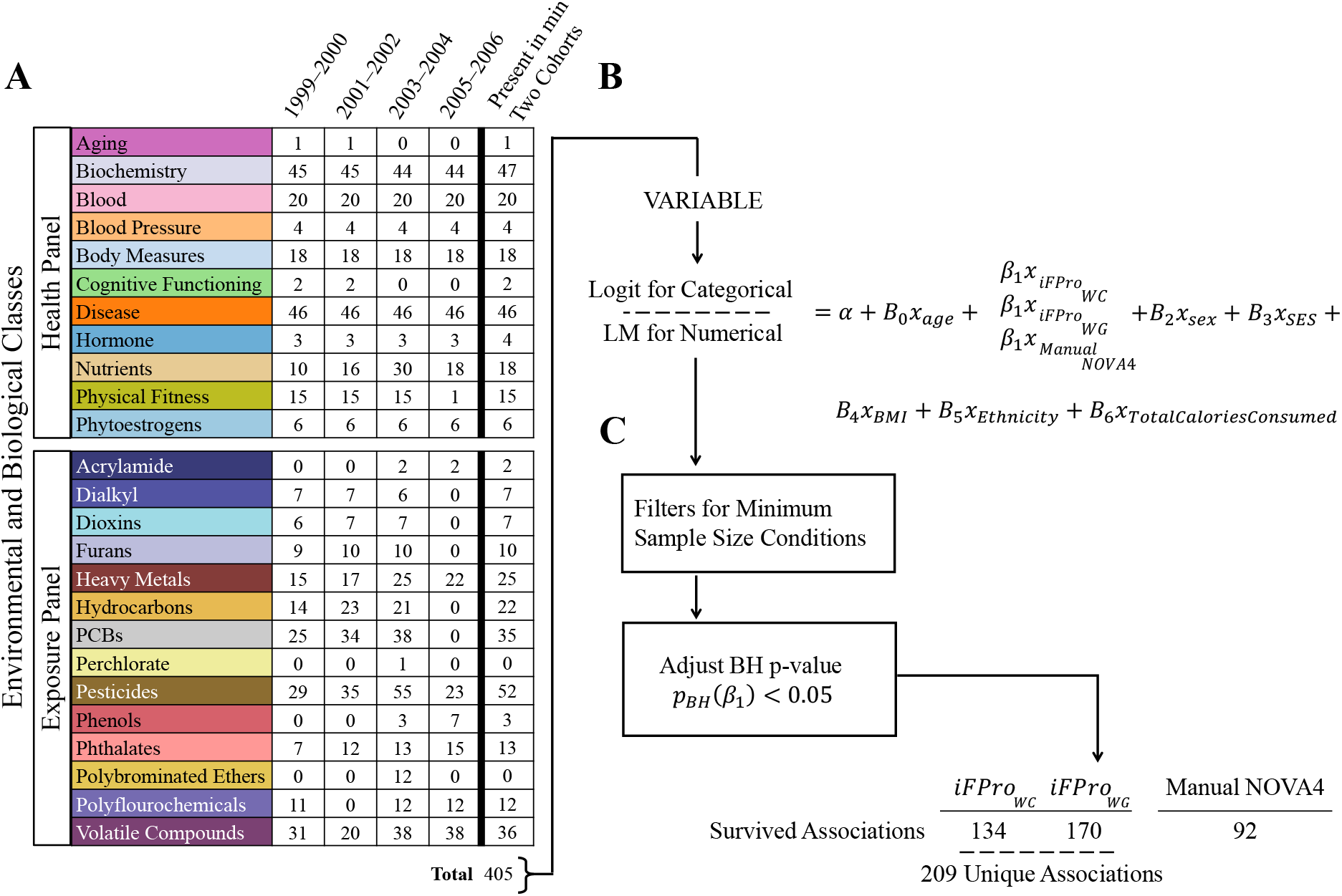
Overview of the EWAS pipeline on NHANES 1999-2006. **(a)** For the selected health phenotypes and chemical exposures provided in each cohort of NHANES, we kept only those variables present in at least two cohorts. **(b)** We investigated possible associations between the selected 405 variables in the combined NHANES 1999-2006 cohort using linear regression for continuous variables and logistic regression for categorical variables. **(c)** To account for false discovery rate, we adjusted the p-value of *β*_1_ using Benjamini-Hochberg method with *α* = 0.05.

All continuous variables were transformed using Box-Cox transformation or logit function (applied to Framingham and ACC/AHA scores) to stabilize the variance and improve the validity of measures of association [30]. We then standardized all continuous variables, to bring their effect sizes on similar scale. For multiple linear regression, we used fully standardized regression coefficients, indicating how many standard deviations of change in the dependent variable are associated with one standard deviation increase in the independent variables. For logistic regression, we opted for a partial standardization, acting only on the continuous independent variables, as we wanted to keep a straightforward interpretation of the relation between one standard deviation increase in the Box-Cox transformed *iFPro* and increase/decrease in disease odds [36].

To account for false discovery rate, we adjusted the p-values corresponding to each score using Benjamini-Hochberg method with *α* = 0.05. Overall, we find 214 significant tests across the three methodologies, with *iFPro*_*W C*_ at 134, *iFPro*_*W G*_ at 170, and manual NOVA 4 at 92. The summary of the analysis is presented in Figures S18-S21. A comparison with literature results based on manual NOVA 4 is reported in Table S9.

**Figure SI. 18:**
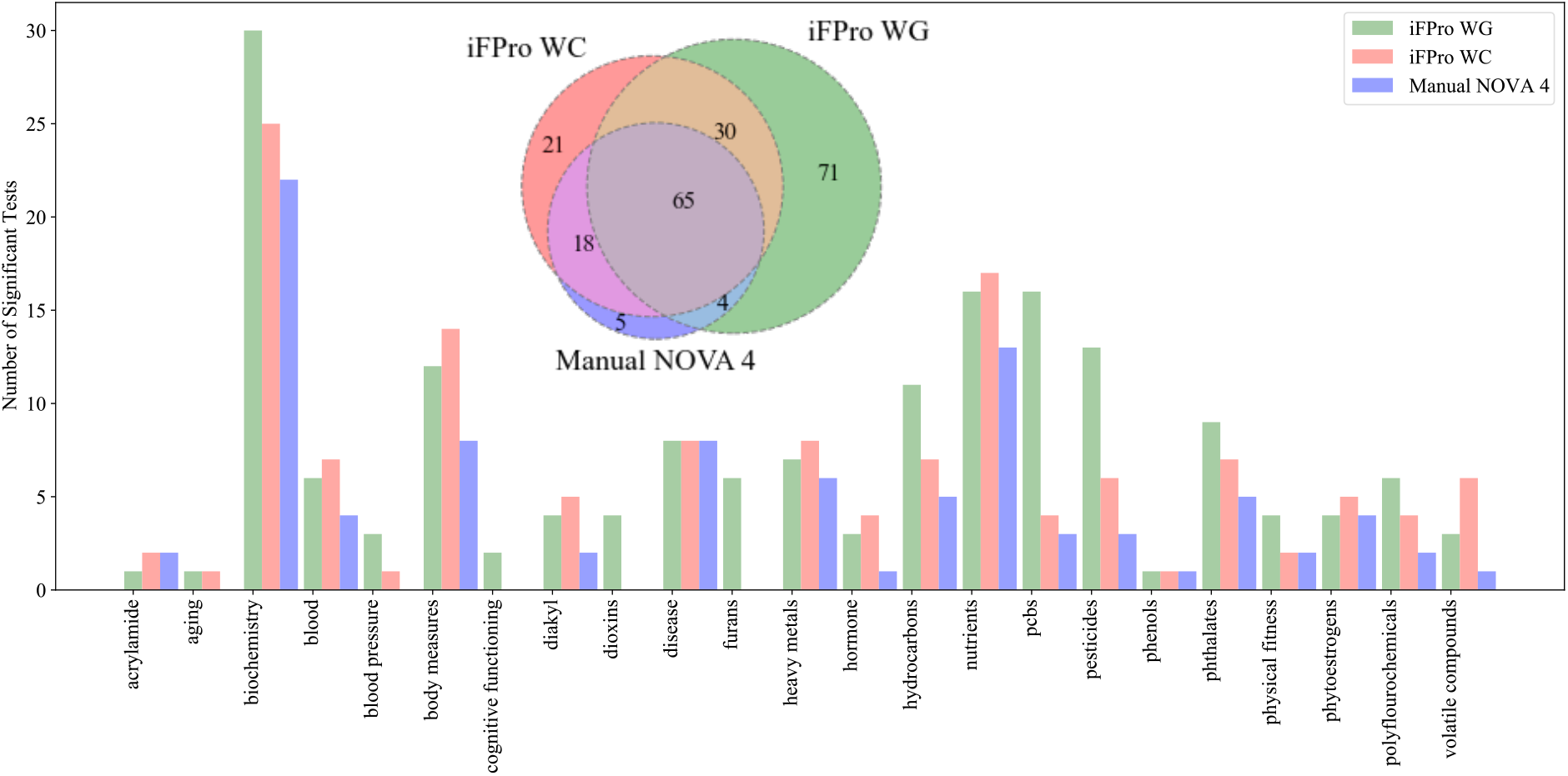
Number of significant tests surviving multiple testing for *iFPro*_*W C*_, *iFPro*_*W G*_, and fraction of caloric intake from manual NOVA 4. For *iFPro*_*W G*_ and *iFPro*_*W C*_, we find 170 and 134 significant associations, respectively, for a total of 209 unique variables. In comparison, the same analysis using the fraction of calories contributed by manual NOVA 4 finds 92 significant associations, of which 95% is in overlap with *iFPro*_*W G*_ and *iFPro*_*W C*_. Overall, the total number of variables recovered by the three methodologies is 214. Here we report the results stratified by module.

**Figure SI. 19:**
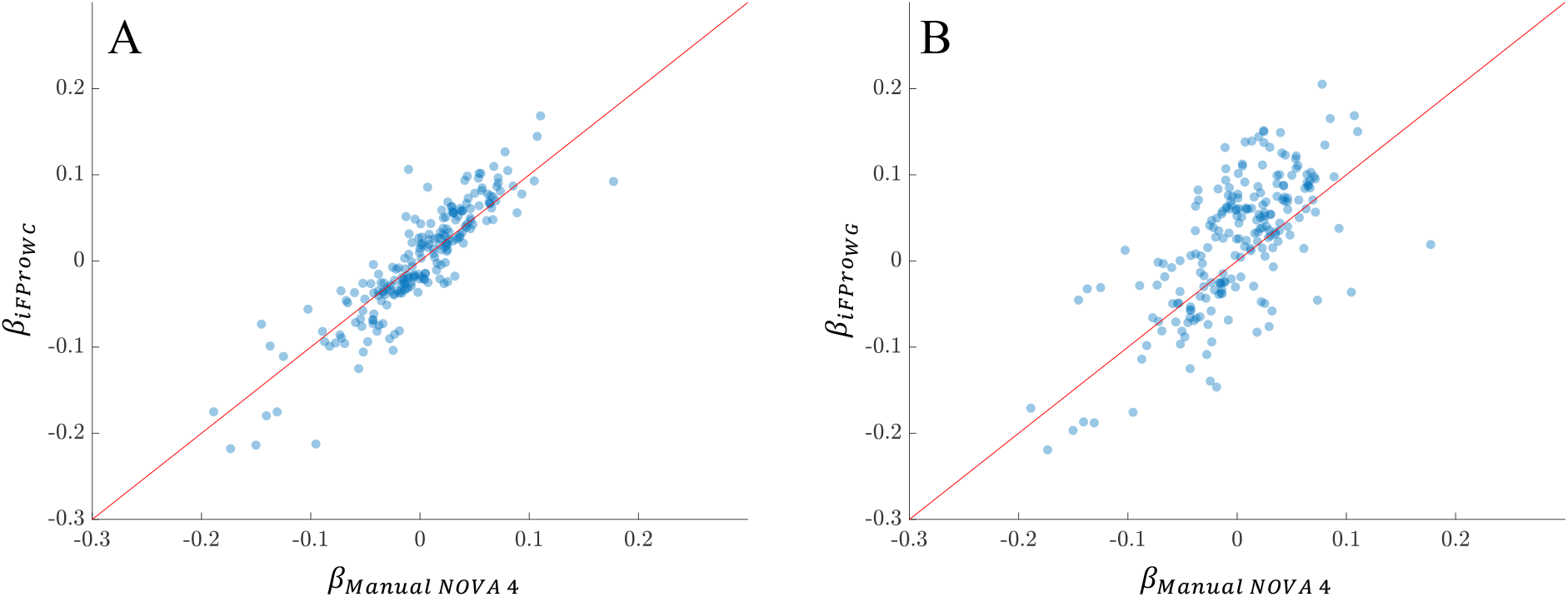
Comparison of the Effect Sizes for the Significant Tests found by *iFPro*_*W C*_, *iFPro*_*W G*_, and fraction of caloric intake from manual NOVA 4. Across the 214 significant variables recovered by the three methodologies we find that **(a)** 70.56% of the times *iFPro*_*W C*_ shows bigger effect sizes compared to manual NOVA 4, while **(b)** for *iFPro*_*W G*_ the percentage increases to 77.57%.

**Figure SI. 20:**
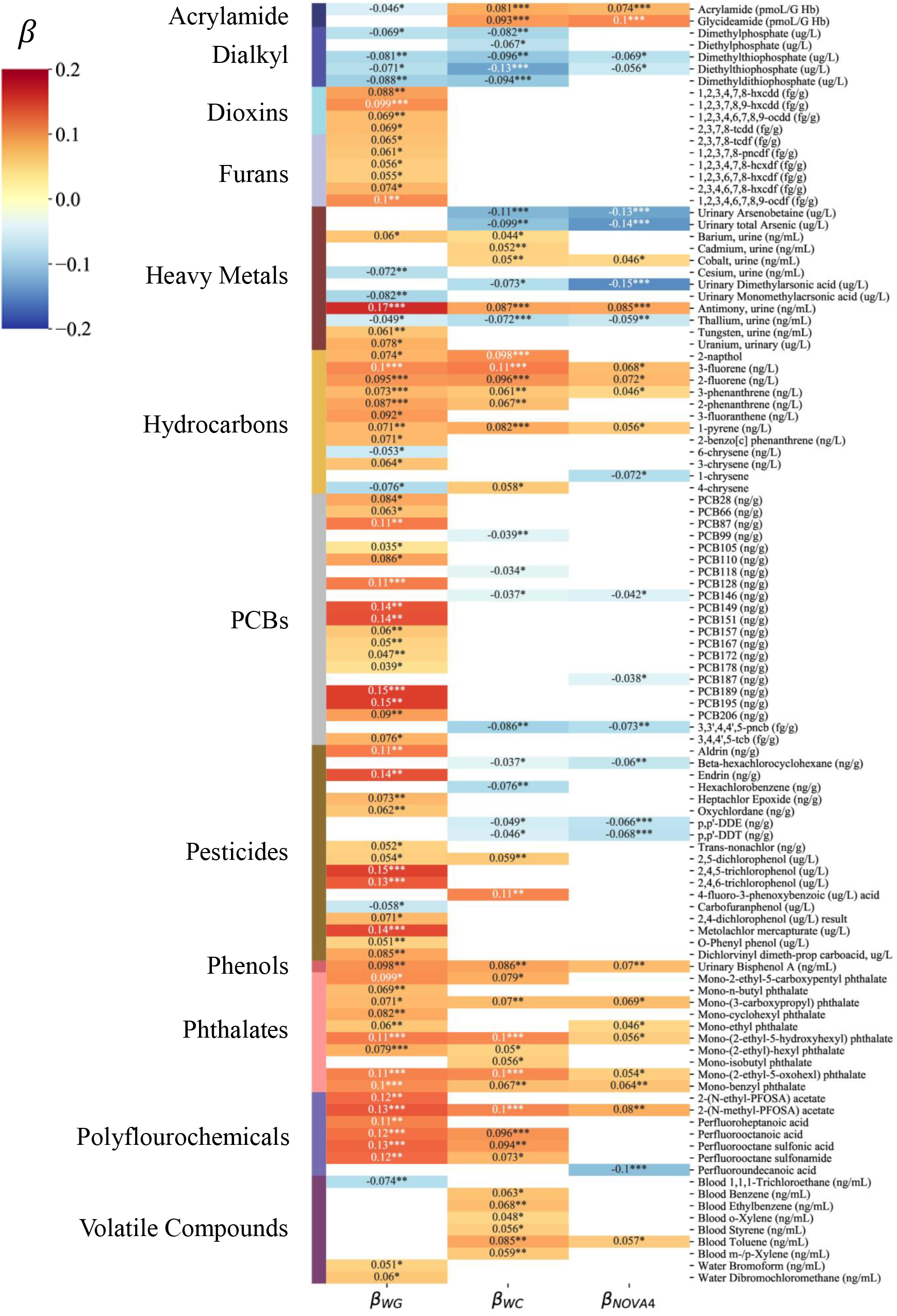
Exposure Panel. Each variable reported on the left (e.g. “Acrylamide”) refers to different exposure modules. We report here the standardized *β* coefficient, quantifying the effect on each exposure when the Box-Cox transformed diet scores increase of one standard deviation over the population. Each variable is color-coded according to *β*, with positive associations in red, and negative associations in blue. In white, we annotate the variables that do not survive Benjamini-Hochberg FDR.

**Figure SI. 21:**
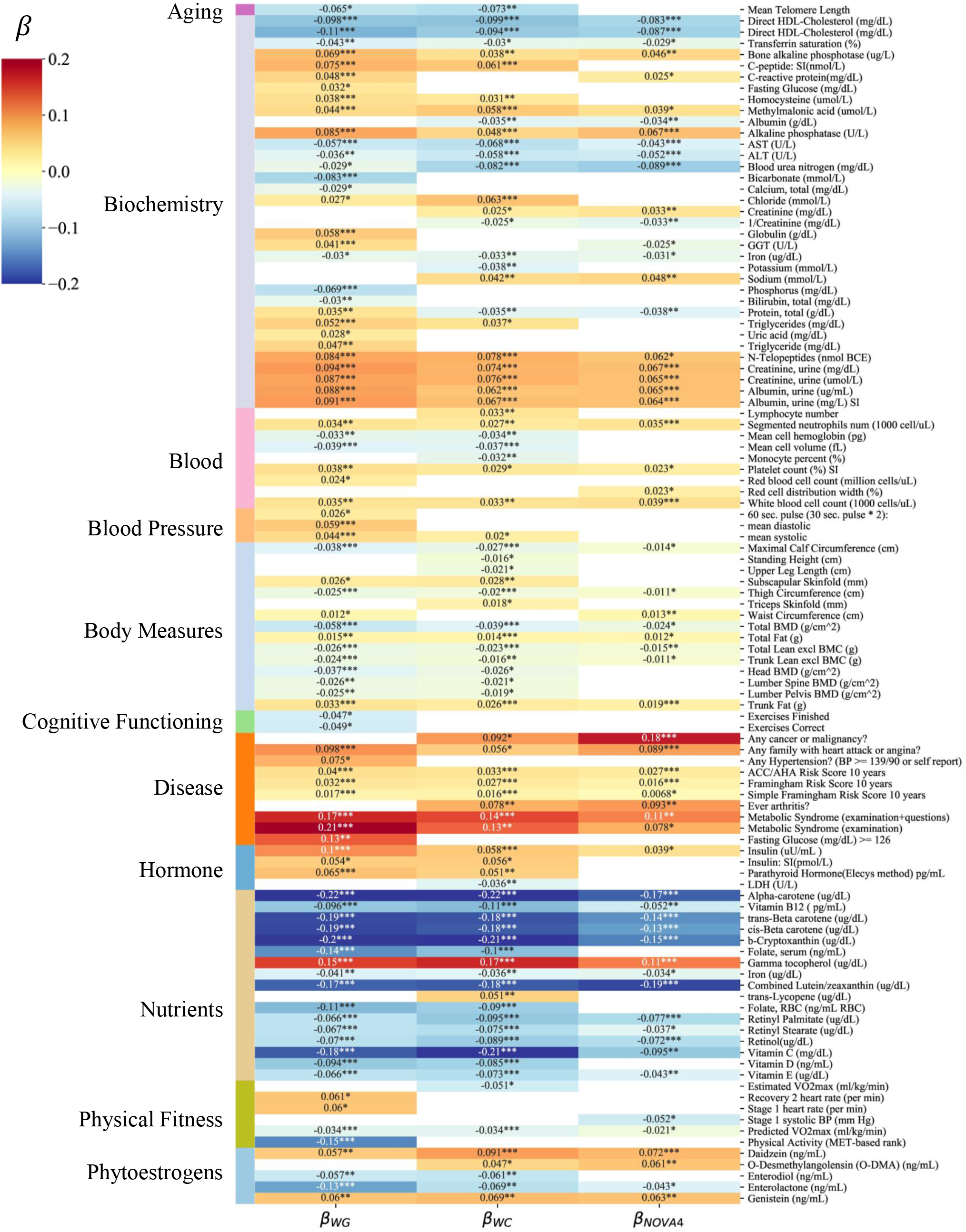
Health Panel. Each variable reported on the left (e.g. “Aging”) refers to different modules of disease phenotypes and measurements assessing the overall health status of each individual. We report here the standardized *β* coefficient, quantifying the effect on each exposure when the Box-Cox transformed diet scores increase of one standard deviation over the population. Each variable is color-coded according to *β*, with positive associations in red, and negative associations in blue. In white, we annotate the variables that do not survive Benjamini-Hochberg FDR.

**Table 9:** A Summary of the Epidemiological Literature on the Discovered Associations with Manual NOVA 4.

| Health or Exposure Panel | Paper | In Agreement with *iFPro* |
| --- | --- | --- |
| Aging | Ultra-processed food consumption and the risk of short telomeres in an elderly population of the Seguimiento Universidad de Navarra (SUN) Project [37] Spain (SUN) | Yes |
| Disease:<br>Cardiovascular | Association between ultraprocessed food intake and cardiovascular health in US adults: a cross-sectional analysis of the NHANES 2011–2016 [38]<br>USA (NHANES)<br>Ultra-processed food intake and risk of cardiovascular disease: prospective cohort study (NutriNet-Santé) [39]<br>France (NutriNet-Santé)<br>Ultra-processed food consumption is associated with increased risk of all-cause and cardiovascular mortality in the Moli-sani Study [40] Molise, Italy (Moli-sani) | Yes |
| Disease:<br>Hypertension | Ultra-processed food consumption and the incidence of hypertension in a mediterranean cohort: The seguimiento universidad de navarra project [41] Spain (SUN) | Yes |
| Continued on next page |  |  |

Table 9 – continued from previous page
| Health or Exposure Panel | Paper | In Agreement with *iFPro* |
| --- | --- | --- |
| Disease:<br>Metabolic Syndrome | Dietary share of ultra-processed foods and metabolic syndrome in the US adult population [42]<br>USA (NHANES)<br>Diet quality indices in relation to metabolic syndrome in an Indigenous Cree (Eeyouch) population in northern Québec, Canada [43]<br>Canada (Aschii Environment & Health Study)<br>A minimally processed dietary pattern is associated with lower odds of metabolic syndrome among Lebanese adults [44] Lebanon | Yes |
| Disease:<br>Cancer | Consumption of ultra-processed foods and cancer risk: results from NutriNet-Santé prospective cohort [45]<br>France (NutriNet-Santé) | Yes |
| Body Measures:<br>Total Fat | Contribution of ultra-processed foods in visceral fat deposition and other adiposity indicators: Prospective analysis nested in the PREDIMED-Plus trial [46]<br>Spain (PREDIMED-Plus) | Yes |
| Continued on next page |  |  |

Table 9 – continued from previous page
| Health or Exposure Panel | Paper | In Agreement with *iFPro* |
| --- | --- | --- |
| Body Measures:<br>Waist Circumference | Ultra-processed food consumption and excess weight among US adults [47] **USA (NHANES)**<br>Consumption of ultra-processed food and obesity: cross sectional results from the Brazilian Longitudinal Study of Adult Health (ELSA-Brasil) cohort (2008–2010) [48] **Brazil (ELSA)**<br>Ultra-processed food consumption and indicators of obesity in the United Kingdom population (2008–2016) [49] **UK (NDNS)** | Yes |
| Nutrients | The share of ultra-processed foods and the overall nutritional quality of diets in the US: evidence from a nationally representative cross-sectional study [50] **USA (NHANES)**<br>Impact of ultra-processed foods on micronutrient content in the Brazilian diet [51] **Brazil (HBS)** | Yes |
| Biochemistry:<br>C-Reactive Protein | Association between consumption of ultra-processed foods and serum C-reactive protein levels: cross-sectional results from the ELSA-Brasil study [52] **Brazil (ELSA)** | Yes |
| Phytoestrogens | Association between dietary share of ultra-processed foods and urinary concentrations of phytoestrogens in the US [53] **USA (NHANES)** | Yes |
| Vegetarian Diet:<br>Adverse Effects of ultra-processing | Consumption of Ultra-Processed Foods by Pesco-Vegetarians, Vegetarians, and Vegans: Associations with Duration and Age at Diet Initiation [54] **France (NutriNet-Santé)** | Yes |

Table 9 – continued from previous page
| Health or Exposure Panel | Paper | In Agreement with *iFPro* |
| --- | --- | --- |
| Phthalates and Phenols | • Ultra-processed food consumption and exposure to phthalates and bisphenols in the US National Health and Nutrition Examination Survey, 2013–2014 [55]<br>USA (NHANES)<br>• Association between dietary contribution of ultra-processed foods and urinary concentrations of phthalates and bisphenol in a nationally representative sample of the US population aged 6 years and older [56]<br>USA (NHANES) | Yes |
| Acrylamides | Association between Heat-Induced Chemical Markers and Ultra-Processed Foods: A Case Study on Breakfast Cereals [57] Spanish supermarkets | Yes |

## 5 Food Substitution

The observed variability of *FPro* for categories of foods similarly consumed in the population (Figure 2E), combined with the EWAS results, quantifying the effect of processed diet on disease risk, suggests a systematic way to implement food substitution and predict its relevance in terms of health indicators [58, 59]. With this goal, we classified all foods consumed by the pooled cohort of 20,047 individuals in NHANES 1999-2006, according to WWEIA [60]. For substitution purposes, the relevance of food *k* in individual *j*’s diet can be quantified as

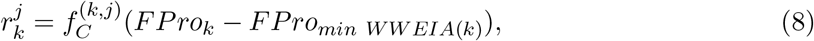

where 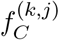 is the fraction of calories contributed by food *k* to the dietary profile, while *FPro*_*min WWEIA*(*k*)_ refers to the food with the lowest *FPro* within the same WWEIA category of food *k*. By picking the suggested foods in the original WWEIA classes reported by each individual, we aim to minimally perturb her habits, to maximize the compliance to the new dietary regime. Moreover, as Eq. S8 offers a heuristic to identify which food to prioritize, the overall level of processing is reduced in a minimal number of steps.

The impact of substituting *M* foods on disease risk is measured in terms of odds ratio (OR), that quantifies the odds of disease occurring when adopting the optimized diet, compared to the original choices. We estimate OR as

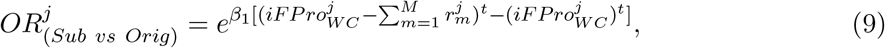

where *β*_1_ is the effect size describing the strength of the association between *iFPro* and disease onset, all 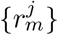 follow from Eq. S8, and with the superscript *t* we denote the function-composition of Box-Cox transformation followed by z-score with parameters estimated in the original population.

For continuous variables *y* like vitamin *B*_12_, vitamin C, and bisphenol A, the impact of substituting *M* foods on individual *j*’s diet is quantified by

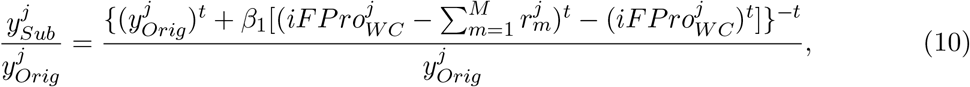

where with the superscript −*t* we refer to the inverse function-composition of Box-Cox transformation followed by standardization (first invert z-score, then Box-Cox).

## 6 Open Food Facts

Open Food Facts is a free, crowd-sourced world-wide database of food products, with nutrition facts and ingredient list [61]. We collected 233,831 nutritional records from their website, corresponding to 168,681 product ids, annotated with NOVA labels according to a heuristic described at [62]. This database also compiled an extensive list of food additives that gave us the possibility to quantify the number of additives per product. Similarly to the cross-validation explained in Section 2.1, we trained and validated two models on the same 5-fold partition: (1) standard FProX leveraging the logarithm of 11 nutrients used as baseline, (2) FProX with an additional input feature capturing the number of additives in each food. The baseline feature panel includes protein, fat, total carbohydrate, sugars, dietary fiber, calcium, iron, sodium, cholesterol, saturated fat, and trans fat. We removed vitamin A and C from the analysis, given the high number of non available values (NAs). Hence, the cross-validation was run on a selection of 228,689 records, with no NAs in any feature.

Overall, we find commendable performances in both scenarios, with higher AUC and AUP when additives are included in the model (see Table 10). In particular, model (2) outperforms model (1) in assessing NOVA 3. We derived *FPro*_1_ and *FPro*_2_ following Eq. 1, finding they highly correlate with each other (*ρ*_*Spearman*_ = 0.9017), and they both correlate well with the number of additives (*ρ*_*Spearman*_(*FPro*_1_, *n*_*additives*_) = 0.6726, *ρ*_*Spearman*_(*FPro*_2_, *n*_*additives*_) = 0.8240). This result is encouraging, as details regarding ingredient list and additives are seldom if ever available in current food composition databases.

To better understand the role of additives in discriminating NOVA classes, we trained and validated a random forest taking the number of additives as the only input, over the same train-test split used for model (1) and (2). We find AUC equal to 0.859923 ± 0.000485 for NOVA 1, 0.832023 ± 0.003375 for NOVA 2, 0.820256 ± 0.000693 for NOVA 3, and 0.907307 ± 0.000609 for NOVA 4. Compared to model (2), the performances in terms of AUC drop of 13.36% for NOVA 1, 15.77% for NOVA 2, 15.03% for NOVA 3, and 7.25% for NOVA 4.

The lack of information on nutrient amounts strongly affect the precision-recall curve, increasing disproportionately the number of false positives predicted for NOVA 1, 2, and 3. Indeed, we find AUP equal to 0.288799 ± 0.000930 for NOVA 1, 0.026227 ± 0.000498 for NOVA 2, 0.439202 ± 0.001269 for NOVA 3, and 0.941548 ± 0.000350 for NOVA 4. The values for class NOVA 1, 2, and 3 are close to the proportion of examples labeled as 1, 2, and 3 in the training, suggesting that the model’s behavior is close to a random classifier. Compared to model (2), the performances in terms of AUP drop of 70.01% for NOVA 1, 97.05% for NOVA 2, 50.96% for NOVA 3, and 4.77% for NOVA 4.

Overall, these results suggest that the number of additives is a good predictor of ultra-processed food as defined by NOVA, but it misses the processing nuances represented in the other NOVA classes.

**Table SI. 10:**
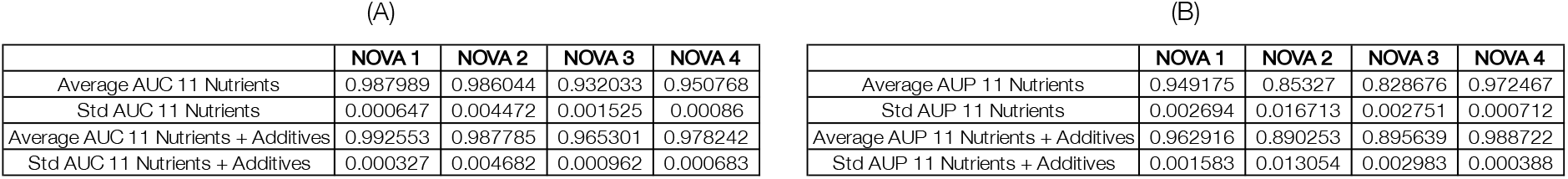
AUC and AUP for the four NOVA classes in Open Food Facts. For model (1) and (2) we report average and standard deviation of AUC (A) and AUP (B) over the stratified 5-folds.

## 7 Data Quality and Future Directions

- **Data Sample and Variability**. The quality of the training data influences our analysis and the interpretation of FPro. Indeed, all machine learning and AI models become feasible only when extensive labeled datasets are available, and on top of that, they provide reliable results when the training data samples the real world in an exhaustive way, avoiding over-fitting.

A single cycle of FNDDS, as well as SR-Legacy, reports representative nutritional average values for each food/drink, which do not capture the variability due to factors such as recipe variations, production methods, soil quality, and storage time. The training data would then benefit of multiple instances of the same food, helping capture the natural variability in nutrient content, and reduce over-fitting [63]. This is why we have investigated how nutritional values for the same food code change through different FNDDS cycles, and how integrating additional variability is affecting FProX and FPro in assessing “unseen foods” (see *Assesment of FPro robustness* below). We envision that Foundation Foods, the new USDA food composition dataset available at FoodData Central, will be a great asset to improve the current data training of FPro, once it will increase its coverage to more than 140 foods. Indeed, Foundation Foods includes individual sample measurements behind the nutrient mean values that populate the other databases, and metadata reporting the number of samples, location, time-stamps, analytical methods used, and, additionally, if available, cultivar and production practices.

A further way to improve how machine learning algorithms and AI characterize food data is to study the statistical properties of nutrient distributions in the food supply, and make sure that the training data sample them exhaustively. In Menichetti et al. [9] we observe how the variability of nutrient concentrations in food, despite covering several order of magnitudes, follows a well-defined functional form, related to the lognormal distribution. This is the reason why we log-transformed all nutrient measures before performing the classification, as this is the natural scale of the nutrient fluctuations. Accounting for the variability driven by chemical reactions in the ingredients, as well as for the variability derived from the complete food production chain, plays a major role in successfully capturing associations between food intake and disease phenotypes [64].

Beyond nutrient variability, the training data would benefit of a better representation of different food groups, as in case of raw meat products, important ingredients in many recipes. This poor representation in the dataset is somehow expected, as FNDDS is designed by the USDA to provide food composition data for foods and beverages reported in the dietary component of NHANES, and raw meat is not commonly consumed by the American population. A better coverage of heterogeneous collection of food groups would also improve the accuracy of the algorithm when facing different national food composition databases, beyond the data collected by the USDA.

Finally, we stress that all potential errors affecting food composition data should generally be random with respect to the machine learning algorithm and the manual NOVA classification that developed FPro, which should only cause attenuation of classification accuracy rather than bias, and attenuation toward the null of all epidemiological outcomes. Thus, our findings would further improve with a more accurate database.

- **Requirements for Branded Products**. While in this study we focus on survey data, our model based on nutritional values can easily work on different food and cohort databases, as proven by our analysis of over 50,000 products collected from major grocery store websites [65]. When facing real-world food data, we have to account for the regulations introduced by government agencies, like the FDA. For instance, in the nutrition facts label nutrients are classified in 3 different classes, characterized by different standards regarding the agreement between declared value on the label and actual values in the sampled food [66]. For nutrients like sugars, total fat, saturated fat, cholesterol, and sodium, the label is considered compliant if the nutrient content of the sampled product is up to 20% above the value declared on the label. Consequently, when relying on the nutrition facts of branded products, FPro should be reported with confidence intervals determined by randomly altering the nutrient content according to FDA regulations.
- **Size and Composition of the Nutrient Panel**. The computation of FPro adapts to different set of nutrients, allowing us to accurately classify food from limited nutrient information. However, the chemical information currently available to train our algorithm is limited by the resolution of food databases. Indeed, many chemicals like acrylamide, ammonium sulfate, azodicarbonamide, butylated hydroxyanisole, and furans, associated with the preparation and preservation of food, are not tracked by national agencies. The lack of quantification of these chemicals becomes even more striking once we acknowledge their impact on human health [67–71]. With a higher number of chemicals available for all foods, we could aim for an unsupervised classification of food processing, eliminating the need for supervised manual curation. Currently, our analysis in Section S1.5 shows that an unsupervised hierarchical clustering of foods, leveraging the widest nutrient panel available in FNDDS, is not able to independently reproduce the four NOVA classes. It is possible, however, that the addition of chemical measurements that pertain to processing signatures could further improve the current results, leading to purely chemically driven classification of food processing.
- **Assessment of FPro robustness**. We aim to test the robustness of FPro by measuring the extent of its variations when re-formulation of the same product are created and measured experimentally. This analysis is currently hindered by limited data availability. However, we were able to test how nutrient variability impacts FPro when comparing the same food code in different cycles of FNDDS. Indeed, while FNDDS does not capture sample variability within the same cycle (e.g. 2009-2010), it documents the nutrient variability affecting the same food through the years, as a function of the specific sample or technique chosen to estimate the representative nutritional average. In FoodProXonFNDDS.xlsx we report the nutrient profile of 5,632 foods, present in both FNDDS 2009-2010 and 2015-2016, and how their FPro changed according to the variations in their nutrient content. For all foods we estimate the number of nutrients that changed between the two editions of FNDDS, accounting for the different rounding approximation affecting each nutrient. In presence of one or more nutrients with maximal variation between 10% and 50%, we observe a median ΔFPro=0.001556 (*Q*_1_=0.000222, *Q*_3_= 0.004764). Allowing significantly bigger fluctuations, as in case of one or more nutrients with maximal variation between 10% and 1000%, we observe a median ΔFPro= 0.003312 (*Q*_1_= 0.000722, *Q*_3_= 0.011310).
- **Limitations of Population Surveys**. To test the prediction power of FPro and iFPro for several health phenotypes and chemical exposures, we leveraged the rich panel of analyses collected in NHANES, a population survey that captures dietary intake with 24-hour recalls. A limitation of the 24-hour recall is its reliance on memory, both for identification of foods eaten as well as for quantification of portion sizes, and the need of highly trained interviewers. Although reliance on the participant’s memory leaves room for measurement error, skilled interviewers can produce highly detailed and useful nutritional data comparable to a dietary record [24]. 24-hour recalls are also affected by random within-person error, typified by the day-to-day fluctuation in dietary intake. Random within-person error tends to decrease correlation and regression coefficients towards zero and to bias relative risks toward one. Multiple days of intake per individual permit an estimate of within-person day-to-day variability, and it is usually statistically more efficient to increase the number of individuals in the sample rather than increase the number of days beyond two per individual [72]. For this reason, NHANES added a second day of dietary intake starting with NHANES 2003 [24]. Despite these limitations, 24-hour recalls are widely used in epidemiology as their statistical issues are well understood, and they were used to rigorously evaluate HEI-2015, a gold-standard non-data-driven dietary score for the epidemiological community [73].

## 8 Overview of FoodProX and FPro

**Figure SI. 22:**
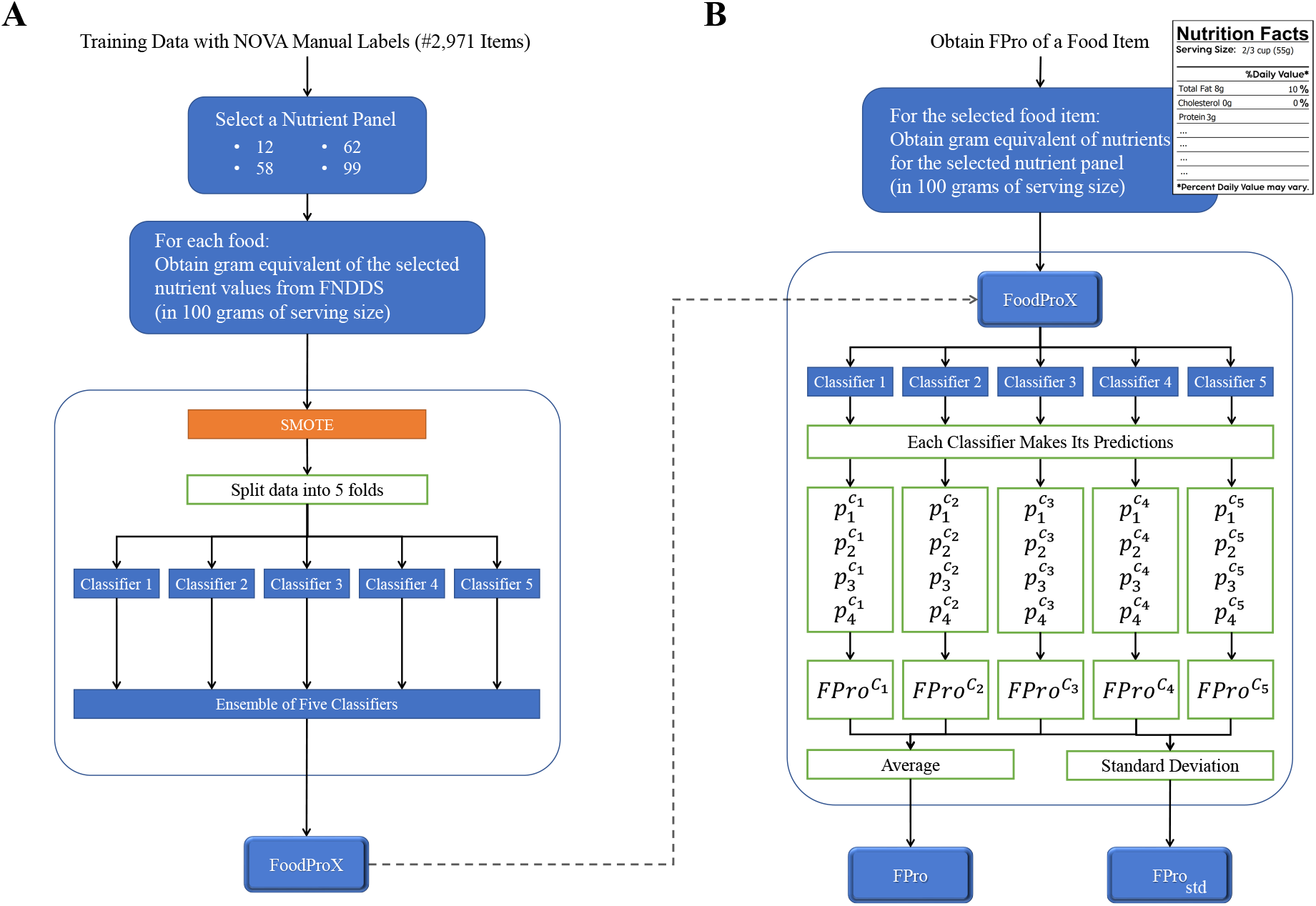
Summary Overview of FoodProX Classifier (A) and FPro Score (B).

